# Autoantibody landscapes in neurological Long COVID and post-COVID cognitive impairment show heterogeneity without a shared disease signature

**DOI:** 10.64898/2026.03.19.26348833

**Authors:** Debanjana Chakravarty, Ravi Dandekar, Vishal D. Lashkari, Iris Tilton, Lindsay McAlpine, Jennifer Chiarella, Allison Nelson, Thomas Ngo, PeiXi Chen, Grace Wang, Aditi Saxena, Bryan Castillo-Rojas, Kelsey Zorn, David R. Tribble, Timothy H. Burgess, Leah H. Rubin, Stephanie A. Richard, Brian K. Agan, Simon D. Pollett, Shelli Farhadian, Serena Spudich, Samuel J. Pleasure, Michael R. Wilson

## Abstract

**Background:** Neurological Long COVID (n-LC) includes persistent cognitive and autonomic symptoms after SARS-CoV-2 infection. Prior studies of post-COVID conditions have described diverse humoral autoreactivity, but findings are heterogeneous, and it remains unclear whether n-LC is associated with a consistent CNS-directed humoral signature.

**Methods:** We performed a cross-cohort case-control analysis to detect autoantibodies in cerebrospinal fluid (CSF) and serum from n-LC participants. In the Yale COVID Mind Study cohort, CSF from n-LC participants and from pre-pandemic and post-COVID asymptomatic controls was assessed by mouse brain immunofluorescence and proteome-wide phage immunoprecipitation sequencing (PhIP-Seq), with candidate reactivities evaluated by orthogonal assays and supervised modeling. In the Epidemiology, Immunology, and Clinical Characteristics of Emerging Infectious Diseases with Pandemic Potential (IDCRP EPICC) cohort, post-COVID sera collected prior to iPhone- or iPad-based cognitive screening were profiled by PhIP-Seq and compared between participants with and without cognitive impairment.

**Results:** CSF immunoreactivity on mouse brain tissue was observed in both n-LC and controls, with similar overall frequencies, although n-LC participants more often showed nuclear-predominant staining patterns. PhIP-Seq identified sparse, largely patient-specific peptide reactivities to nuclear and neuronal proteins in CSF and serum. Supervised models provided limited discrimination between cases and controls. Candidate autoantigens had limited disease specificity on orthogonal testing. EPICC serum autoantibody profiling similarly failed to distinguish individuals with and without cognitive impairment.

**Conclusions:** Across cohorts and compartments, n-LC did not exhibit a shared autoantibody signature. These findings support the absence of a dominant, common CNS autoantibody-mediated mechanism in n-LC.

**Funding:** Grants HU00012020067, HU00012120103, HU00011920111, R01NS125693, R01MH125737, R01AI157488 from Defense health program and NIH.

## Introduction

Approximately 10-30% of individuals infected with SARS-CoV-2 experience a wide range of persistent post-COVID symptoms lasting for months to even years, affecting one or more organ systems and collectively referred to as Long COVID (LC) (1–7). Among the LC population, significant numbers of patients report neurologic symptoms, including headaches, fatigue, cognitive difficulties often characterized as ‘brain fog’, and autonomic nervous system dysfunction (8–13). Despite affecting millions worldwide, the biological underpinnings of neurologic LC (n-LC) remain elusive.

Several non-exclusive mechanisms have been proposed. Persistent viral reservoirs or residual viral antigens may sustain a low-level systemic immune activation, including the development of pathogenic autoantibodies targeting the central nervous system (CNS). This is largely based on studies of acute SARS-CoV-2 infection, which revealed profound immune dysregulation, with transient loss of immune tolerance and the development of de novo autoantibodies directed against nuclear, phospholipid, cytokine, and cell-surface antigens (14–22). Although these responses typically wane after recovery from the acute viral infection, several studies have reported persistent autoreactivity months later, correlating with ongoing symptoms (23–27). Most LC studies identify at least one class of autoantibody associated with disease persistence, though findings vary across cohorts and methodologies. Particularly, antinuclear antibodies (ANAs), which are the hallmarks of systemic autoimmunity, have been reported at higher titers or prevalence in several LC cohorts compared with fully recovered individuals, sometimes correlating with fatigue or dyspnea (26–29). Alongside these systemic markers, functional autoantibodies against G-protein-coupled receptors (GPCRs), including β₂-adrenergic, muscarinic, and angiotensin II receptors, have been detected in subsets of patients with fatigue, cognitive, and autonomic dysfunction (30–36); however, one of these studies did not have any control comparison group (31). Another study using rapid extracellular antigen profiling (REAP) identified individual autoantibodies in LC patients, but no significant differences were noted in the number of autoreactivities observed between LC vs recovered controls (37).

Furthermore, evidence from cerebrospinal fluid (CSF) studies illustrates similar uncertainty. In a study of 50 individuals with self-reported cognitive complications 8 months post-infection, 52% had anti-neuronal antibodies in serum but only 6% in CSF. Notably, there was no control group, and analyses relied on post-mortem tissue (38). Another study showed normal CSF parameters and no recognizable anti-neuronal autoantibodies in the CSF of the majority of the patients. Only 12% of patient sera exhibited anti-myelin autoantibodies (39). Despite these inconsistent CSF findings, passive-transfer experiments provide a complementary line of evidence: IgG purified from individuals with LC can induce pain hypersensitivity, balance disturbances, persistent sensory hypersensitivity, and reduced locomotor activity in animal models, suggesting that patient autoantibodies can be biologically active and potentially pathogenic in a subset of cases (40, 41). Moreover, nearly all prior studies have relied on targeted assays, such as ELISAs, microarrays, or cell-based systems, to detect known antigens. These methods are powerful when a candidate antigen is predefined, but inherently limited in scope if autoreactivity in n-LC targets unrecognized neural proteins.

To address these limitations, we used an unbiased proteomic discovery tool called phage immunoprecipitation sequencing (PhIP-Seq) (42, 43) to identify autoantibodies in the CSF and serum, utilizing two different independent cohorts of individuals with n-LC. The first cohort, the Yale COVID Mind Study (44), enrolled participants with n-LC, control participants who recovered from COVID-19 without neurologic sequelae, and pre-pandemic controls. The second cohort, the Infectious Disease Clinical Research Program- Epidemiology, Immunology, and Clinical Characteristics of Emerging Infectious Diseases with Pandemic Potential (IDCRP EPICC) cohort (45, 46), enrolled LC participants who underwent iPhone/iPad-based cognitive testing and were stratified by cognitive impairment status. Through this multi-cohort design, we sought to define whether a convergent autoantibody signature underlies n-LC.

## Results

### Cohort characteristics and group-level demographic/clinical summaries

Cohort demographics and clinical features are summarized in Table 1, and the enrollment and experimental workflow are outlined in Figure 1. The Yale COVID Mind Study prospectively enrolled participants and collected CSF with paired serum from n-LC (n=31), post-COVID asymptomatic controls (n=7), and pre-pandemic Never-COVID controls (n=22) (Figure 1a). An independent EPICC cohort (n=73) was stratified by Brain-Baseline Assessment of Cognition and Everyday Functioning (BRACE) screening into cognitively impaired (BRACE+, n=29) vs unimpaired (BRACE-, n=44); serum was collected approximately 7 months after COVID-19 (IQR 6-12) and before BRACE measurement, with additional pre-pandemic (n=12) and healthy donor sera (n=40) as controls (Figure 1b).

**Figure 1.**
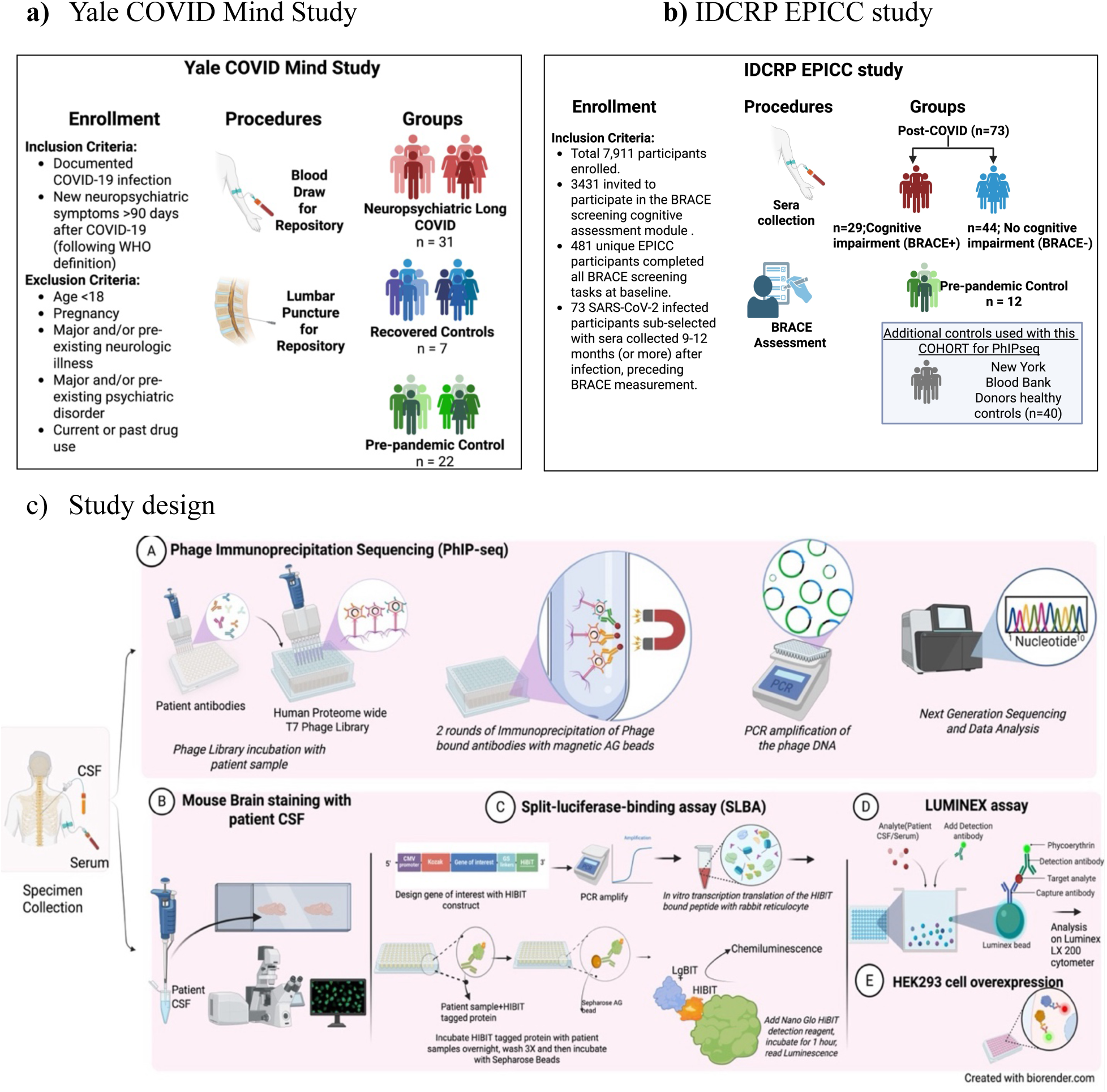
a) Patient data collection summary a) Yale COVID Mind Study, b) IDCRP EPICC study (c) Schematic of the study design. CSF supernatant and serum samples from were used for different downstream analyses. A) In the Phage Immunoprecipitation Sequencing (PhIP-Seq) assay, a T7 human proteome phage display library was incubated with patient samples. Antibody-bound phages were captured using magnetic AG beads. The DNA was amplified and sequenced to identify specific antibody targets. B) Mouse anatomical brain staining was performed with fixed mouse brain sections, incubated and stained with patient CSF, and visualized using fluorescence microscopy. C) The split-luciferase binding assay (SLBA) was used to orthogonally validate the PhIP-Seq peptides. Peptides tagged with HIBIT were incubated with patient samples, and luminescence was measured. D) Luminex bead-based assay was used to quantify SARS-CoV-2 antibodies in CSF and serum. E) Another way of orthogonal validation, the PhIP-Seq peptide of interest was overexpressed in HEK293 cells and incubated with patient CSF.

**Table 1.** Demographic and clinical characteristics of participants in The COVID Mind Study at Yale and the IDCRP EPICC cohort.

|  |  | The COVID Mind Study at Yale Cohort |  |  |  | EPICC Cohort |  |  |
| --- | --- | --- | --- | --- | --- | --- | --- | --- |
|  |  | Never COVID Control (n=22) | Neurologic Long COVID (n=31) | Post-COVID Asymptomatic (n=7) | p value (Yale) | BRACE Not Impaired (N = 44) | BRACE Impaired (N = 29) | p value (EPICC) |
| Age | Median [IQR] | 47.5 [33 – 55] | 53 [40 – 61] | 41 [31 – 48] | 0.2603 | 43 [36.5 – 53] | 38 [30 – 50] | 0.3462 |
|  | Mean ± SD | 45.6 ± 13.16 | 49.5 ± 12.92 | 40.4 ± 13.67 |  | 44.8 ± 11.09 | 41.7 ± 15.06 |  |
| Sex | Female | 6 (27.3%) | 21 (67.7%) | 5 (71.4%) | 0.0076 | 14 (32%) | 18 (62%) | 0.0158 |
|  | Male | 16 (72.7%) | 10 (32.3%) | 2 (28.6%) |  | 30 (68%) | 11 (38%) |  |
| Active Duty | Yes | - | - | - |  | 24 (54.5%) | 14 (48.3%) | 0.6386 |
| Race & Ethnicity | White | 7 (31.8%) | 25 (80.6%) | 3 (42.9%) | 0.0009 | 31 (70.5%) | 19 (65.5%) | 0.0811 |
|  | Black/African American | 10 (45.5%) | 5 (16.1%) | 2 (28.6%) |  | 1 (2%) | 6 (21%) |  |
|  | Hispanic | 4 (18.2%) | 1 (3.2%) | 0 (0.0%) |  | 7 (16%) | 3 (10%) |  |
|  | Asian | 1 (4.5%) | 0 (0.0%) | 2 (28.6%) |  | 3 (7%) | 1 (3%) |  |
|  | Multiple | - | - | - |  | 2 (4.5%) | 0 (0%) |  |
| BMI | Median [IQR] | 27.3 [25.5-31.70] | 28.8 [25.52-31.78] | 25.5 [23.2-27.0] | 0.6257 | 30.2 [27.0 – 34.2] | 27.4 [24.1 – 32.2] | 0.1455 |
|  | Mean ± SD | 28.2 ± 4.73 | 29.8 ± 7.2 | 27.9 ± 9.8 |  | 30.4 ± 5.56 | 28.4 ± 5.53 |  |
| Days between COVID-19 illness and blood draw | Median [IQR] | - | 319 [243-436] | 323 [137-413] |  | 202.0 [188.5, 360.2] | 253.0 [193.0, 376.0] |  |
|  | Range | - | 68-460 | 81-701 |  | 167-588 | 169-585 |  |
| Acute COVID-19 Level of Care | Home (n, %) | - | 21 (67.7%) | 6 (85.7%) | 0.1486 | 35 (79.5%) | 27 (93.1%) | 0.2972 |
|  | Hospital Floor (n, %) | - | 6 (19.4%) | 0 (0.0%) |  | 8 (18.2%) | 2 (6.9%) |  |
|  | ICU (n, %) | - | 4 (12.9%) | 0 (0.0%) |  | - | - |  |
|  | Unknown (n, %) | - | 0 (0.0%) | 1 (14.3%) |  | - | - |  |
|  | Asymptomatic | - | - | - |  | 1 (2.3%) | 0 (0) |  |
| Vaccination Status | Fully vaccinated | - | 3 (9.7%) | 3 (42.9%) | 5.94E-06 | 35 (79.5%) | 24 (82.8%) | 1.0000 |
|  | Partially vaccinated | - | 0 (0.0%) | 1 (14.3%) |  | - | - |  |
|  | Unvaccinated | - | 28 (90.3%) | 0 (0.0%) |  | 9 (20.5%) | 5 (17.2%) |  |
|  | Unknown | - | 0 (0.0%) | 3 (42.9%) |  | - | - |  |
| Co-morbidities | Smoking (current or former) | 14 (63.6%) | 10 (32.3%) | 2 (28.6%) | 0.0683 | 18/42 (42.9%) | 3/27 (11.1%) | 0.0069 |
|  | Alcohol use (excluding the unknown status) | 9 (40.9%) | 0 (0.0%) | 0 (0.0%) | 5.52E-05 | 8/42 (19%) | 6/27 (22%) | 0.7672 |
|  | Hypertension | 2 (9.1%) | 6 (19.4%) | 1 (14.3%) | 0.6830 | 10/41 (24%) | 4/27 (15%) | 0.3790 |
|  | Type 2 Diabetes (T2D) | 3 (13.6%) | 3 (9.7%) | 0 (0.0%) | 0.7096 | 3 (7%) | 2 (7%) | 1.0000 |
|  | Obesity | 6 (27.3%) | 9 (29.0%) | 1 (14.3%) | 0.8429 | 22 (50%) | 10/27 (37%) | 0.3325 |
|  | Antidepressant use (12 mo) | 0 (0.0%) | 2 (6.5%) | 2 (28.6%) | 0.0489 | Not collected | Not collected |  |
| CCI Score | Median [IQR] | - | - | - |  | 0.00 [0.00, 1.00] | 0.00 [0.00, 1.00] |  |
|  | Range | - | - | - |  | 0.00 – 6.00 | 0.00 – 9.00 |  |
| Other | PROMIS cognitive impaired (n = 60) | - | - | - |  | 10/38 (26%) | 7/22 (32%) | 0.7681 |
|  | DoDSR (pre-COVID) sera specimen present | - | - | - |  | 8 (18%) | 5 (17%) | 1.0000 |
| Neuro Long COVID symptoms | Cognitive dysfunction | - | 25 (81%) | - |  | - | - |  |
|  | Fatigue/Post exertion malaise (PEM) | - | 24 (77%) | - |  | - | - |  |
|  | Mood changes | - | 24 (77%) | - |  | - | - |  |
|  | Headaches | - | 18 (58%) | - |  | - | - |  |
|  | Neuropathy | - | 17 (55%) | - |  | - | - |  |
|  | Dizziness/Falls | - | 17 (55%) | - |  | - | - |  |
|  | Dysautonomia | - | 15 (48%) | - |  | - | - |  |
|  | Poor sleep | - | 12 (39%) | - |  | - | - |  |
| CCI = Charlson Comorbidity Index; PROMIS = Patient Reported Outcomes Measurement System; DoDSR - Department of Defense Sera Repository |  |  |  |  |  |  |  |  |
| BRACE = Brain Baseline Assessment of Cognition and Everyday Functioning |  |  |  |  |  |  |  |  |
| EPICC = Epidemiology, Immunology, and Clinical Characteristics of Emerging Infectious Diseases With Pandemic Potential |  |  |  |  |  |  |  |  |
| - Data Not available |  |  |  |  |  |  |  |  |
| P values for categorical variables were calculated using exact tests (Fisher's exact test for 2x2 tables and Fisher-Freeman-Halton exact tests for larger contingency tables). |  |  |  |  |  |  |  |  |
| Continuous variables were calculated using Welch's ANOVA for Yale and Welch's two-sample t-test for EPICC cohort. |  |  |  |  |  |  |  |  |

In the Yale cohort, sex distribution differed across groups (p=0.0076), with n-LC (21/31; 67.7% female) and post-COVID asymptomatic controls (5/7; 71.4% female) enriched for females compared with Never-COVID controls (6/22; 27.3% female) (Table 1). Race/ethnicity also differed (p=0.0009). n-LC participants were predominantly White (25/31; 80.6%), with a smaller representation of Black/African American (5/31; 16.1%) and Hispanic (1/31; 3.2%). Never-COVID controls included a higher proportion of Black/African American (10/22; 45.5%) and Hispanic (4/22; 18.2%) participants and a lower proportion of White (8/22; 36.4%) (Table 1). Among reported covariates, alcohol use (excluding the unknown status) differed across Yale groups (p=5.52e-05), reported in 9/22 (40.9%) Never-COVID controls and 0/7 (0%) post-COVID asymptomatic controls but 0/31 (0%) n-LC participants, and antidepressant use in the prior 12 months also differed (p=0.0306) (Table 1). Vaccination status differed between n-LC and post-COVID asymptomatic controls (p= 5.94e-06): most n-LC participants were unvaccinated (28/31; 90.3%), whereas post-COVID asymptomatic controls included 3/7 (42.9%) fully vaccinated, 1/7 (14.3%) partially vaccinated, and 3/7 (42.9%) with unknown status. Within the Yale n-LC group, the most commonly reported LC symptoms were cognitive dysfunction (25/31, 81%), fatigue/post-exertional malaise (24/31, 77%), mood changes (24/31, 77%), headaches (18/31, 58%), neuropathy (17/31, 55%), dizziness/falls (17/31, 55%), dysautonomia (15/31, 48%), and poor sleep (12/31, 39%) (Table 1).

In EPICC, BRACE- and BRACE+ participants were similar in overall cohort structure but differed in select characteristics. Both groups were predominantly identified as white (BRACE- 31/44; 70.5%; BRACE+ 19/29; 65.5%) and largely fully vaccinated (BRACE- 35/44; 79.5%; BRACE+ 24/29; 82.8%). Serum was collected prior to BRACE testing, at a median of 202.0 days from infection to blood draw in BRACE- and 253.0 days in BRACE+. Sex distribution differed by BRACE status (p=0.0158), with BRACE+ participants more often female (18/29; 62%) compared to BRACE- participants (14/44; 32%). Smoking history also differed (p=0.0069), with 18/42 (42.9%) BRACE- participants reporting it compared with 3/27 (11.1%) BRACE+ participants.

### Tissue-based immunofluorescence revealed similar frequencies and brain-region reactivity between n-LC cases, pre-pandemic, and post-COVID asymptomatic controls

Mouse brain sections were immunostained using diluted CSF (1:10) from participants within the Yale COVID Mind study cohort. Positive staining was observed in 9 of 31 n-LC samples (29%), 6 of 22 Never-COVID controls (27%), and 1 of 7 post-COVID controls (14%). Fisher’s exact test showed no significant difference in staining frequency between groups (p = 0.84, ns) (Figure 2a).

**Figure 2.**
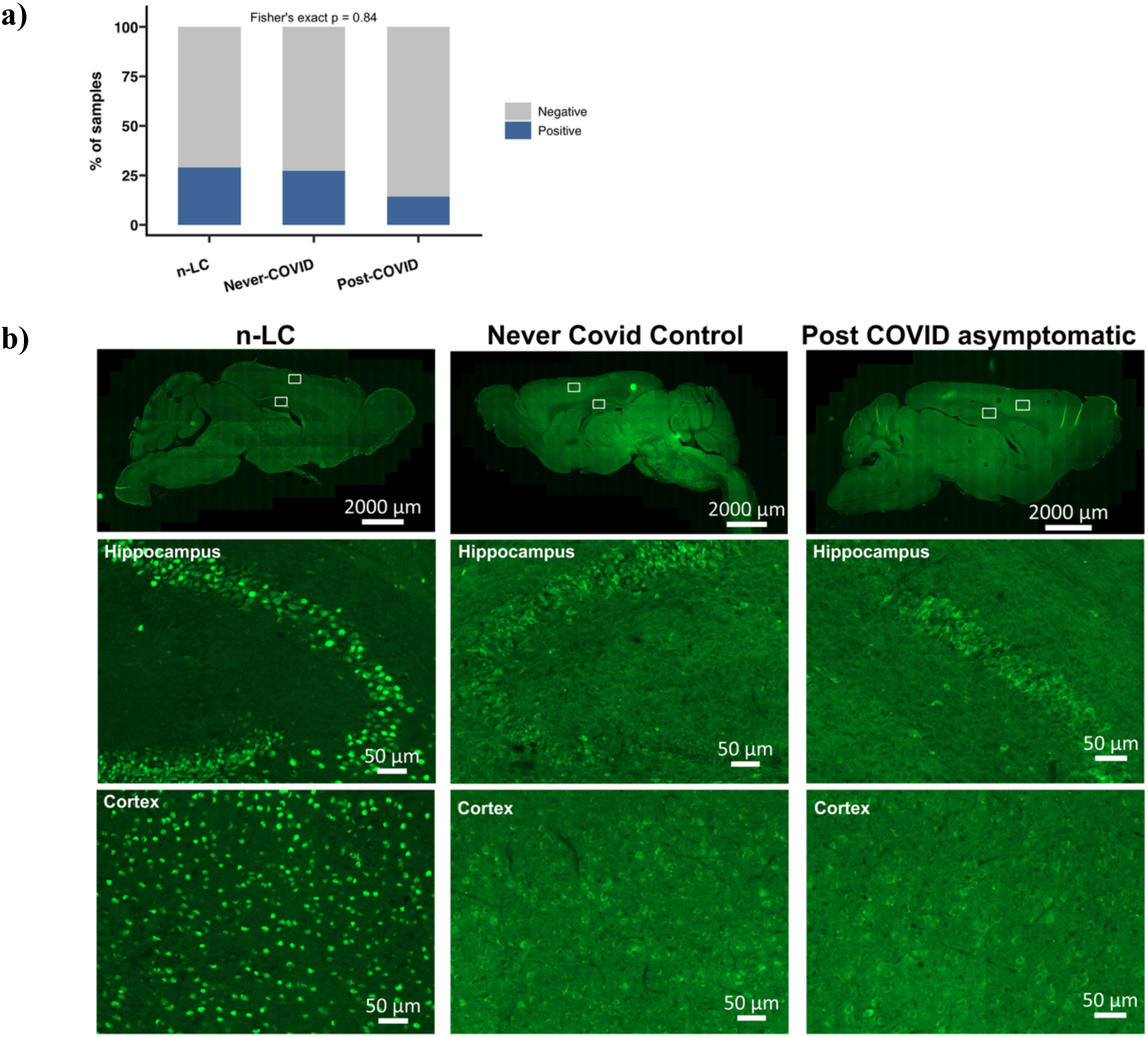
Tissue-based immunofluorescence revealed broadly similar frequency and brain region reactivity between neurological Long-COVID (n-LC) cases vs pre-pandemic (Never-COVID) controls and post-COVID asymptomatic controls but showed distinct cellular staining patterns. Mouse brain sections were immunostained using CSF from neurological Long-COVID (n-LC, n=31), pre-pandemic Never-COVID controls (n=22), and post-COVID asymptomatic controls (n=7) and visualized with anti-human IgG to screen for anti-neural autoantibodies. a) Staining was observed in 9/31 n-LC (29%), 6/22 Never-COVID (27.3%), and 1/7 post-COVID (14.2%) controls. Fisher’s exact test showed no significant difference in the staining frequency between groups (p = 0.84, ns). b) Nuclear-dominant patterns were more frequent in n-LC cases, while majority of the controls exhibited weaker cytoplasmic staining as shown in the representative whole-brain scans (top; scale bar, 2,000µm) and their boxed regions corresponding to higher-magnification fields in cerebellum, and hippocampus (scale bar, 50um).

All staining-positive samples were re-stained at a 1:4 dilution to enhance signal intensity and enable more detailed assessment of regional, cellular, and subcellular patterns (Figure 2b). The hippocampus, thalamus, hypothalamus, Purkinje cells, and brainstem are the prominent sites of reactivity across groups, including Never-COVID controls, post-COVID controls, and n-LC cases. In both control groups, immunoreactivity was predominantly cytoplasmic, with a few samples showing cytonuclear staining, largely restricted to gray matter, with minimal white matter staining. Purkinje cells were consistently positive, and neuropil labeling was frequently observed (Figure 2b). A small subset of controls showed an interneuron-like cytoplasmic pattern distributed broadly across the brain, along with fiber-like staining in the cortex and cerebellum. A greater proportion of n-LC cases exhibited stronger nuclear-predominant staining, often with little or no cytoplasmic or neuropil labeling. One n-LC case (case_Y10) displayed a distinct phenotype with intense cytoplasmic staining in the nucleus accumbens and deep cerebellar nuclei, together with robust neuropil staining in the dentate gyrus and extending mossy fibers (Supplementary Figure 1a). Overall, staining phenotypes among n-LC cases included strong nuclear staining in 5/9, cytonuclear staining in 3/9, and cytoplasmic-only staining in 1/9. Figure 2b highlights representative differences in cerebellar and hippocampal staining between n-LC cases and controls. Collectively, immunostaining revealed heterogeneous antibody-binding patterns across all groups, with no difference in staining frequency between n-LC and controls.

### PhIP-Seq revealed heterogeneous CSF autoreactivities from n-LC cases showing positive brain immunostaining

To investigate whether the neuronal immunostaining in n-LC corresponded to specific molecular targets, PhIP-Seq was performed on CSF from patients with n-LC (n = 31) and pre-pandemic Never-COVID controls (n = 22). Within the n-LC group, samples were stratified by their mouse brain staining phenotype into stain-positive (n-LC+; n = 9) and stain-negative (n-LC-; n = 22) sub-groups.

There was heterogeneous peptide enrichment across samples, with no single peptide persistently enriched in all, or even most, positively stained n-LC samples (Figure 3a). Additionally, the total number of enriched peptides per sample did not differ significantly between positively and negatively stained n-LC cases (Mann-Whitney test, p = 0.47, ns; Figure 3b).

**Figure 3.**
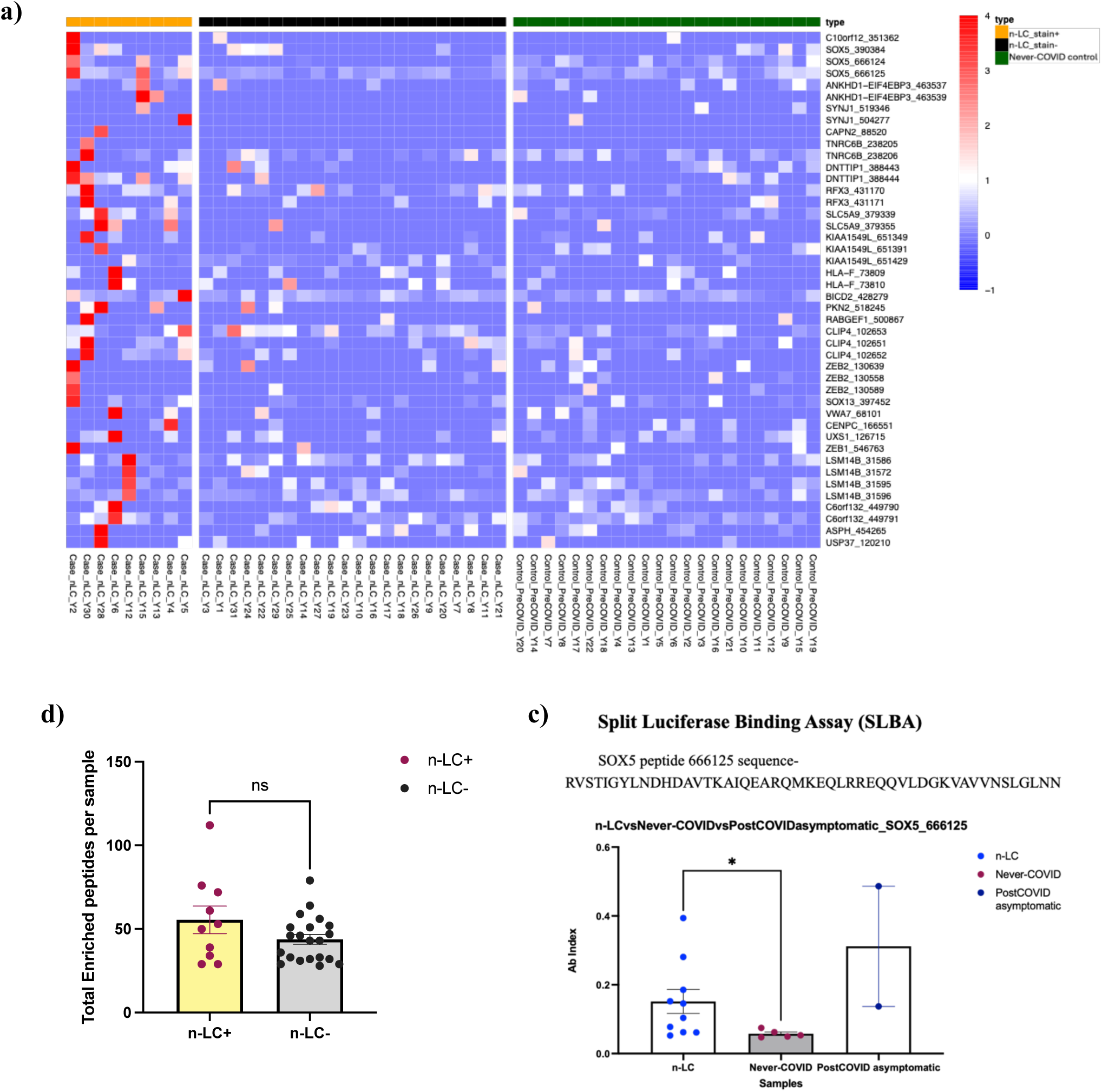
a) Heatmap of human PhIP-Seq reactivity in n-LC CSF samples stratified by mouse anatomical brain staining status and compared to all Never-COVID controls. This heatmap displays peptide-level enrichment data from human phage immunoprecipitation sequencing (PhIP-Seq) performed on cerebrospinal fluid (CSF) samples from patients with neurological Long-COVID (n-LC) and pre-pandemic healthy controls (Never-COVID). n-LC samples are stratified based on prior staining on mouse brain sections. These are compared to CSF samples from Never-COVID controls. Each row corresponds to a unique peptide, and each column represents an individual CSF sample. Color intensity reflects the degree of peptide enrichment, quantified as the log₁₀-transformed fold changes in cases relative to controls. **b) No significant difference was observed in the total number of enriched peptides per sample between the positive and negatively stained n-LC group**. Comparisons between two groups of samples were performed using the Mann-Whitney test. **c) Orthogonal validation of Sox5 (SRY-box transcription factor 5) by Split Luciferase Binding assay (SLBA).** Dot blots show SOX5 peptide 666125 SLBA enrichments (antibody indices) in randomly selected n-LC patients (n = 10; including the 2 patients that enriched the peptide on PhIP-Seq) relative to randomly chosen Never-COVID controls (n = 5) and post-COVID asymptomatic controls (n=2). The experiment was performed in duplicate for each sample. Comparisons between two groups of samples were performed using a two-tailed Mann-Whitney test.

Enriched peptides belonged to nuclear transcriptional factors and chromatin-associated proteins such as SRY-box transcription factor 5 (SOX5), Regulatory Factor X3 (RFX3), Deoxynucleotidyltransferase Terminal Interacting Protein 1 (DNTTIP1), and DEAD-Box Helicase 24 (DDX24), alongside synaptic and neural membrane proteins including Synaptojanin-1 (SYNJ1), Adhesion G Protein-Coupled Receptor V1 (ADGRV1), and voltage-gated potassium channel (KCNQ4). Additional candidate autoantigens involved cytoskeletal/endosomal trafficking peptides and extracellular matrix peptides. However, each peptide was only enriched by 1 or 2 patient CSFs out of the total 9 n-LC patients that stained positively in mouse anatomical brain staining. Two overlapping SOX5 peptides were detected in 2 independent participants. SOX5 is a transcriptional regulator with critical roles in neuronal differentiation and CNS development and has been implicated in systemic autoimmune conditions such as rheumatoid arthritis and psoriatic arthritis (47). It has established roles in Th17 cell differentiation and IL-6-mediated signaling, as well as recent evidence of autoreactive B-cell activation and immune dysregulation in COVID-19, linking SOX5 expression to CD11c⁺/T-bet⁺ atypical memory B cells (48). Moreover, SOX5 genetic variants have been identified within susceptibility networks for fatigue-dominant LC (49). Based on these converging findings, SOX5 was prioritized for further validation.

### SOX5 orthogonal validation on SLBA showed limited specificity for n-LC

SOX5 reactivity in the CSF was tested using SLBA directed at peptide index 666125 (peptide sequence RVSTIGYLNDHDAVTKAIQEARQMKEQLRREQQVLDGKVAVVNSLGLNN) from the peptidome phage library. Antibody indices in randomly selected n-LC patients (n = 10, including the only two CSF samples with enrichment on the PhIP-Seq assay) were compared to Never-COVID (n = 5) and post-COVID asymptomatic control CSFs (n = 2) (Figure 3c). A SOX-5 antibody index was significantly different in n-LC cases compared to Never-COVID controls (p=0.0280, two-tailed Mann-Whitney test). However, no difference was observed between n-LC cases and post-COVID asymptomatic controls. Hence, SOX5 reactivity may reflect a broader post-infectious immune activation, but it was not unique to n-LC.

### Cohort-wide (non-staining-stratified) PhIP-Seq and Logistic Regression analyses showed limited case-control discrimination and heterogeneous CSF autoreactivities in n-LC

Since mouse brain immunostaining did not show a significant difference in staining frequency between n-LC cases and controls, we next asked whether CSF from individuals with n-LC harbored autoantibody profiles distinct from those of controls, regardless of staining results on mouse brain sections. A Logistic Regresion (LR) analysis was performed using peptide-level enrichment data (Methods). The classifier showed only moderate discrimination between all n-LC CSF samples (n = 31) and pre-pandemic Never-COVID controls (n = 22), with the best model achieving an AUC of 0.74, the average AUC across 100 iterations = 0.63, and the lowest AUC = 0.51 (Figure 4a). Among the top features contributing to case-control classification, several peptides corresponded to transcriptional regulatory proteins, cytoskeletal trafficking, and membrane proteins with established CNS expression and neuronal roles (Figure 4b).

**Figure 4.**
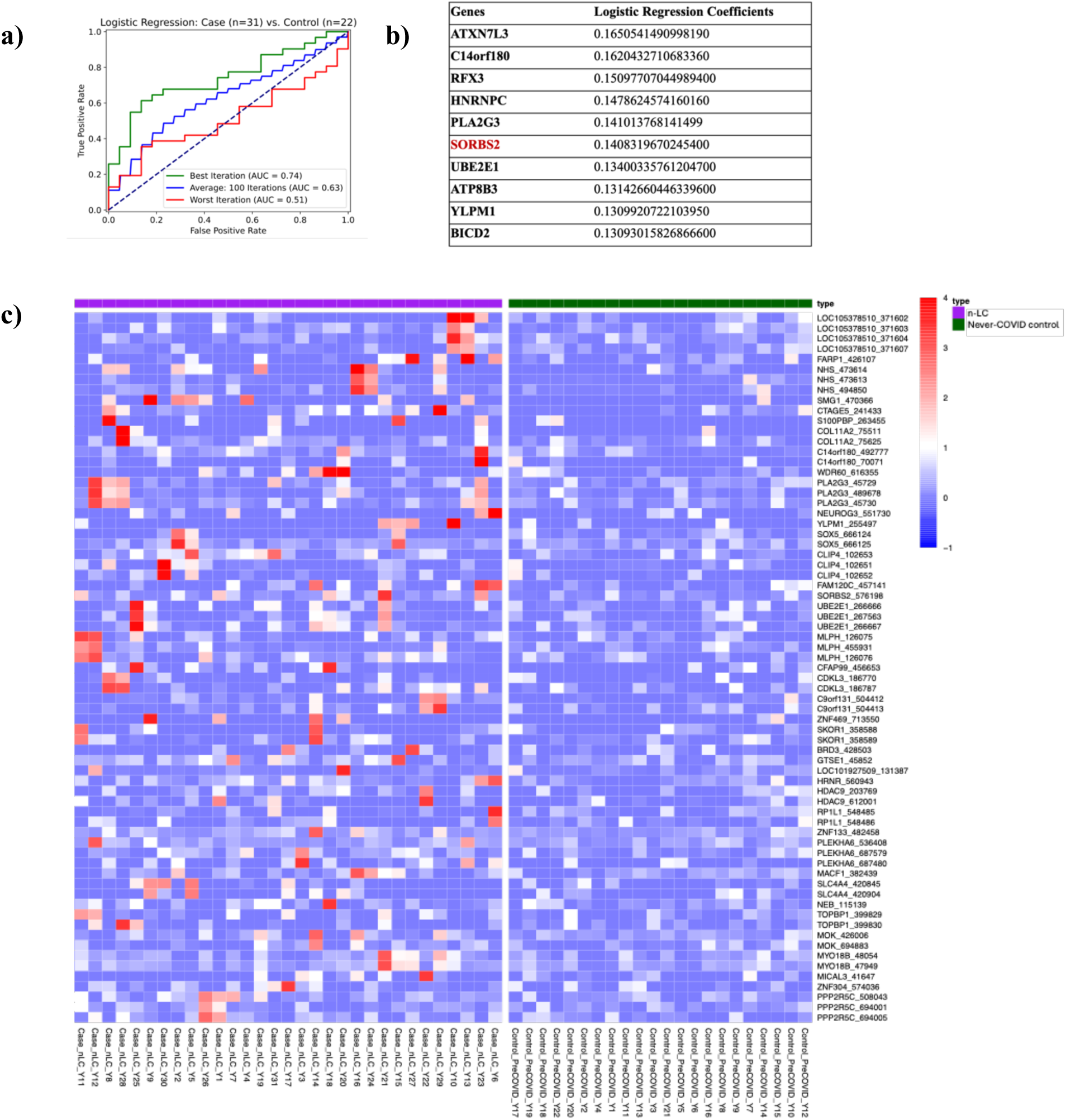
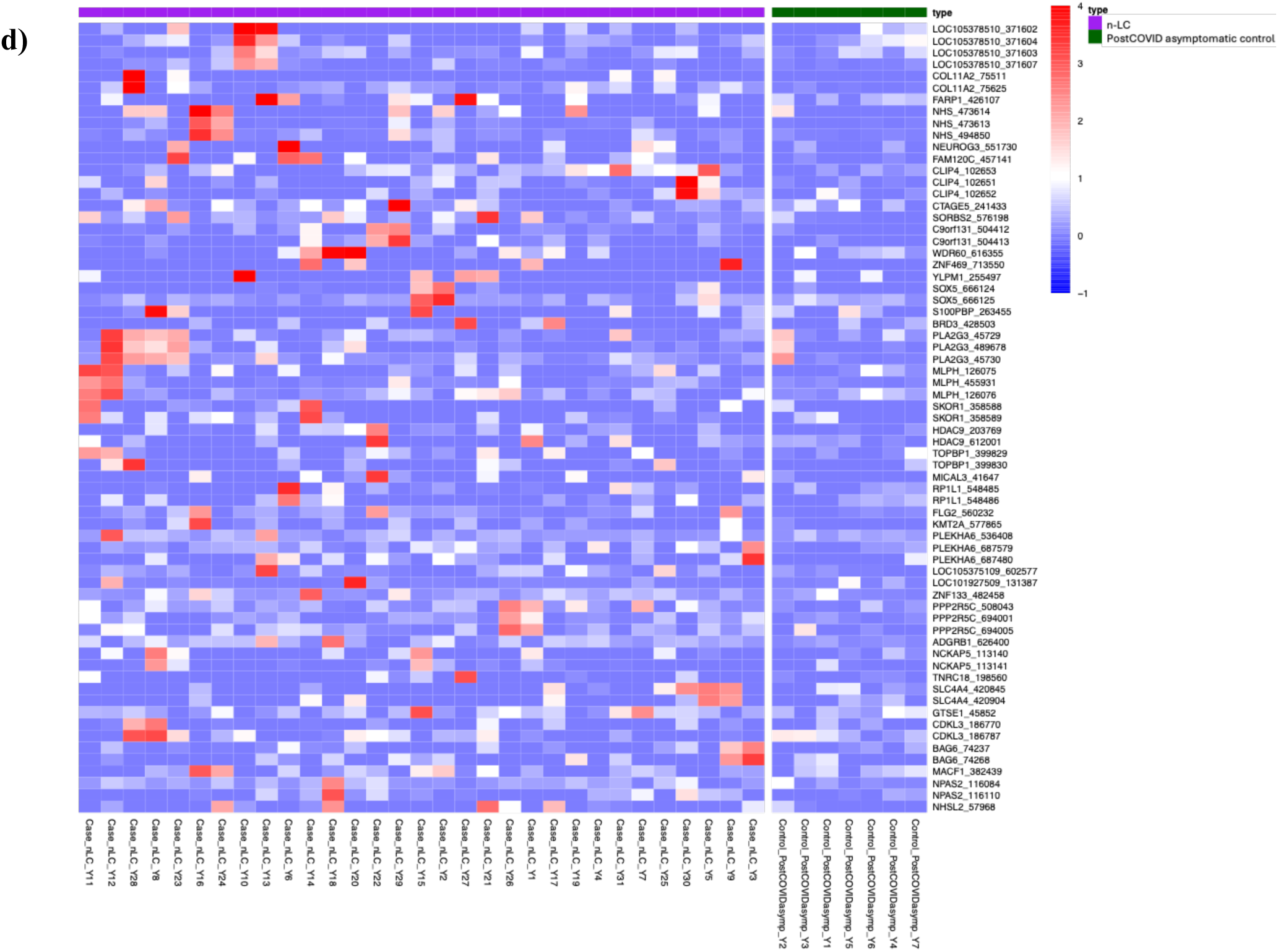
a) Logistic regression classifier shows a modest performance in distinguishing between all n-LC CSF samples (n = 31) from all Never-COVID Control CSF (n = 22). ROC curves from logistic regression models across 100 iterations. The green curve represents the best iteration with the highest area under the curve (AUC = 0.74), indicating moderate discriminatory ability. The blue curve shows the average performance across all iterations (AUC = 0.63), and the red curve indicates the worst iteration (AUC = 0.51), reflecting near-random classification. **b) Table showing the top 10 genes ranked by their logistic regression coefficients.** These genes contribute the most to case-control classification. **c)Heatmap of human PhIP-Seq reactivity in all n-LC CSF samples compared to all Never-COVID controls.** This heatmap displays peptide-level enrichment data from human phage immunoprecipitation sequencing (PhIP-Seq) performed on cerebrospinal fluid (CSF) samples from all patients with neurological Long-COVID (n-LC) (irrespective of their staining pattern) and pre-pandemic healthy controls (Never-COVID). Each row corresponds to a unique peptide, and each column represents an individual CSF sample. Color intensity reflects the degree of peptide enrichment, quantified as the log₁₀-transformed fold changes in cases compared to controls. **d) Heatmap of human PhIP-Seq reactivity in all n-LC CSF samples compared to post-COVID asymptomatic controls**. This heatmap displays peptide-level enrichment data from human phage immunoprecipitation sequencing (PhIP-Seq) performed on cerebrospinal fluid (CSF) samples from all patients with neurological Long-COVID (n-LC) and post-COVID asymptomatic controls. Each row corresponds to a unique peptide, and each column represents an individual CSF sample. Color intensity reflects the degree of peptide enrichment, quantified as the log₁₀-transformed fold change relative to controls.

Consistent with this poor discrimination between groups, PhIP-Seq heatmaps comparing n-LC CSF to Never-COVID controls (Figure 4c) demonstrated highly variable peptide-level enrichment. Enriched peptides were not shared among more than 2-4 n-LC participants, representing ∼6-13% of the total n-LC patients. A similar pattern was observed when comparing n-LC CSF to post-COVID asymptomatic controls (Figure 4d), again revealing low-frequency, individualized reactivities rather than a unifying autoantigenic signature.

Given that the SORBS2 peptide was one of the top peptides in LR, and it recurrently enriched across both n-LC versus Never-COVID and n-LC versus post-COVID CSF comparisons, we sought to validate this candidate by CBA (SORBS2 plasmid -pcDNA3.1-SORBS2-C-(K)-DYK (Genscript, OHu19421D). SORBS2 is a synaptic regulatory protein highly expressed in the hippocampus and implicated in dendritic spine growth, maturation, and excitatory synaptic transmission (50, 51). HEK293T cells transfected with FLAG-tagged SORBS2 were probed with n-LC CSF samples and visualized with anti-human IgG secondary antibody. No colocalization was observed between CSF immunoreactivity and SORBS2 overexpression (Supplementary Figure 1b).

Furthermore, a reciprocal PhIP-Seq analysis using control samples as reference was performed to assess potential background reactivity. When Never-COVID control CSF was compared to n-LC cases (Supplementary Figure 2), peptides that surpassed the enrichment thresholds were only detected in one or two individuals at most. None overlapped with peptides enriched in n-LC samples. A similar result was obtained when using post-COVID asymptomatic CSF as the reference (Supplementary Figure 3), which again revealed enrichments in one or two individuals. Notably, one post-COVID asymptomatic control also enriched a SORBS2 peptide, but the peptide corresponded to a different region of the protein than that recognized by n-LC cases. Collectively, these analyses confirm that control CSF samples exhibit only sporadic, individualized autoreactivities, and the diversity seen in n-LC reflects patient-specific immune repertoires.

### Serum PhIP-Seq profiling revealed heterogeneous and low-frequency antibody reactivities similar to CSF

To determine whether the antibody reactivities observed in CSF were mirrored in the periphery, antibody repertoires were examined in the paired serum samples. Serum samples from n-LC participants were compared to pre-pandemic Never-COVID controls (Supplementary Figure 4). Consistent with CSF findings, no single peptide or protein target was commonly enriched across participants. Peptides meeting enrichment thresholds were detected in only 2-4 n-LC samples. The top enriched peptides included SOX5, Nuclear Factor of Activated T-cells 4 (NFATC4), Rho Guanine Nucleotide Exchange Factor 17 (ARHGEF17), and Bromodomain-containing protein 1 (BRD1), representing proteins involved in neuronal differentiation, cytoskeletal organization, and signal transduction. The presence of overlapping hits such as SOX5 in both CSF and serum datasets suggests systemic rather than compartmentalized autoreactivity, yet these signals remained low-frequency and non-convergent across the cohort.

### SARS-CoV-2-specific antibody responses in CSF and serum were comparable across n-LC and control participants

Antibody index values were derived from CSF and serum MFI and total IgG (Methods). Across all groups, log Antibody Index was not significantly different for Spike or Nucleocapsid (Kruskal-Wallis test; ns) (Supplementary Figure 5). These findings demonstrate comparable SARS-CoV-2-specific antibody responses in n-LC cases and controls, indicating that compartmentalized antiviral antibody production is unlikely to underlie n-LC.

### Autoantibody profiling in the EPICC cohort revealed limited discrimination between post-COVID participants with and without cognitive impairment

A LR classifier trained on peptide-level PhIP-Seq enrichment features showed limited ability to distinguish BRACE+ from BRACE- sera in the independent EPICC cohort, with the best iteration achieving an AUC of 0.61, the mean AUC across 100 iterations = 0.51, and the worst iteration AUC = 0.39 (Figure 5a). Overall performance remained close to 0.5, indicating minimal predictive power of serum PhIP-Seq reactivity patterns to separate individuals with cognitive impairment from those without in this cohort. PhIP-Seq heatmaps comparing BRACE+ and BRACE- sera relative to controls similarly revealed sparse, heterogeneous, and low-frequency peptide enrichment, without a consistent reactivity signature across BRACE+ cases (Figure 5b).

**Figure 5.**
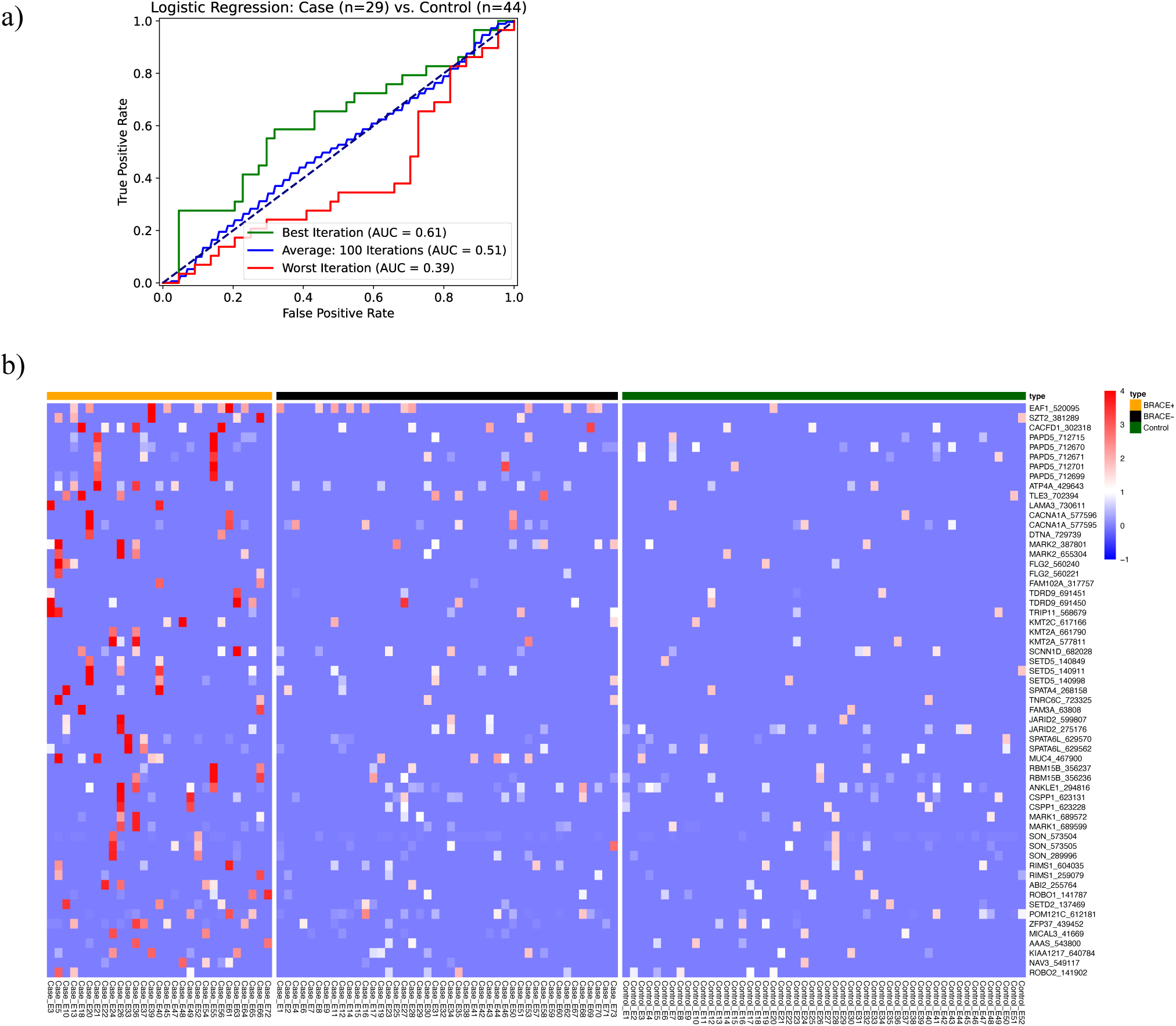
a) Logistic regression classifier could not distinguish between all BRACE+ post-COVID serum (n = 29) from BRACE- post-COVID serum (n = 44) from the EPICC study. This plot indicates little to no predictive power, with best iteration achieving the AUC of 0.61, the average across 100 iterations 0.51, and the worst 0.39. **b) Heatmap of human PhIP-Seq reactivity in BRACE+ post-COVID serum cases compared to BRACE- post-COVID serum cases and controls.** This heatmap displays peptide-level enrichment data from human phage immunoprecipitation sequencing (PhIP-Seq) performed on serum samples from patients with objective cognitive dysfunction versus patients without cognitive impairment, normalized above controls. Each row corresponds to a unique peptide, and each column represents an individual serum sample. Color intensity reflects the degree of peptide enrichment, quantified as the log₁₀-transformed fold changes in cases relative to controls.

Several peptides derived from CNS-related proteins were enriched in two or three BRACE+ participants at most (Figure 5b), including CACNA1A (P/Q-type Ca²⁺ channel controlling synaptic release), SZT2 (mTOR-pathway regulator of neuronal excitability), ROBO2 (axon-guidance receptor essential for cortical connectivity), DTNA (postsynaptic cytoskeletal adaptor), MARK2 (microtubule-associated kinase directing neuronal polarity), along with additional nuclear transcription factors and non-neuronal peptides.

In fact, when all post-COVID serum samples from participants (BRACE+ and BRACE-) were analyzed alongside pre-pandemic and healthy donor controls, LR showed similarly weak discrimination (best AUC = 0.62). Only ∼4-14 % of cases shared an enriched peptide, reinforcing the absence of a shared post-COVID autoantibody signature (Supplementary Figure 6).

## Discussion

This study provides a comprehensive assessment of CNS humoral autoimmunity in n-LC. Across tissue-based assays and peptide-level, whole-human-proteome PhIP-Seq profiling, the data consistently reveal a lack of convergent, disease-specific autoantibody signatures.

Although a subset of n-LC samples demonstrated immunoreactivity in rodent brain tissue, neither the staining frequency nor the major brain regions involved differed significantly from those in the control groups. Also, the staining-positive n-LC subgroup itself was phenotypically heterogeneous, comprising nuclear-predominant, cytonuclear, and cytoplasmic-only patterns rather than a single recurring subcellular staining profile. Taken together, these data do not support an n-LC-specific CNS immunostaining signature at the group level but instead indicate heterogeneous antibody-binding phenotypes within and across groups.

Human proteome-wide PhIP-Seq profiling further supported this heterogeneity. Among CSF samples, enriched peptides consisted of transcriptional regulators (SOX5, RFX3, DDX24), chromatin-associated proteins (DNTTIP1), and neuronal or synaptic proteins (SYNJ1, ADGRV1, KCNQ4). However, enrichment was highly individualized, and no single peptide was enriched across more than two positively stained n-LC patients. Modest SOX5 reactivity was confirmed in a subset of n-LC samples. However, the lack of difference between n-LC and post-COVID asymptomatic controls suggests that SOX5 reactivity only reflects an immune activation state post virus infection rather than being a specific hallmark of n-LC.

Furthermore, logistic regression analyses reinforced these observations. The overall n-LC cohort could only be moderately discriminated from pre-pandemic controls based on PhIP-Seq peptide enrichments. Candidate genes contributing most strongly to classification, such as SORBS2, BICD cargo adaptor 2 (BICD2), and RFX3, are proteins with well-defined neuronal or cytoskeletal roles, but SORBS2 did not validate as an autoantigen by CBA. Meanwhile, reciprocal analyses using control CSF as the reference showed similarly sparse but non-overlapping enrichments, highlighting that low-frequency reactivities occur sporadically in both cases and controls. Interestingly, a single post-COVID asymptomatic control exhibited SORBS2 reactivity, but to a distinct peptide segment from that enriched in n-LC cases, underscoring that minor autoreactivities can emerge stochastically across individuals. Serum analyses paralleled the CSF findings. Enriched peptides again occurred in only a minority of n-LC participants. Overlapping targets between compartments, such as SOX5 suggest that some autoreactivities may reflect systemic immune activation rather than CNS-restricted phenomena. SARS-CoV-2-specific spike and nucleocapsid antibody titers in CSF and serum were comparable between n-LC and control groups, suggesting that there was no ongoing, compartmentalized anti-viral antibody response in the CNS.

A second independent cohort (EPICC) aimed at understanding the association between autoimmunity and objective cognitive impairment (BRACE+) yielded similar results. LR failed to distinguish BRACE+ from BRACE- participants, and only 4-14 % of post-COVID sera showed any peptide enrichment above controls, which is similar to the 6-12% of n-LC cases (in both CSF and serum) that showed autoreactivities above controls. While some CNS-linked peptides, such as CACNA1A, ROBO2, DTNA, and RIMS1, were identified, they were only enriched in a limited number of individuals, reinforcing the absence of a coherent autoantibody signature underlying cognitive dysfunction. Notably, although GPCR-class autoantibodies are prominently observed to be associated with LC(31, 32), as discussed previously, our datasets (both cohorts) showed only sparse GPCR-related signals, including adhesion GPCRs and GPCR-trafficking proteins such as ADGRB1 and GPRASP1, and Rho GTPase/cytoskeletal regulators like ARHGAP21 and ARHGEF17, without convergence at the group level. Ubiquitin- and transcription-related targets also appeared as individualized findings in our cohort. So, a class-level overlap exists (GPCR-related, nuclear/transcriptional, ubiquitin/proteostasis), even when specific antigens differ across studies. Along with our findings, another published PhIP-Seq study of post-COVID cohorts likewise reported no distinct serum autoantibody profile distinguishing LC from recovered individuals, but showed differences compared to uninfected controls (52). Despite a lack of a unifying association with LC in our and other studies, changes in host autoantibody profiles after SARS-CoV-2 infection have been noted in other longitudinal cohorts. For example, new-onset autoantibodies emerged and persisted after SARS-CoV-2 infection, with most (73%) directed at intracellular proteins; however, there was extreme inter-individual variability (24).

Taken together, this study does not support the hypothesis that n-LC and post-COVID cognitive syndromes are characterized by a unifying autoantibody repertoire. Instead, the humoral responses observed likely represent residual immune activation following infection or baseline antibody reactivity rather than an active autoimmune process. These findings suggest that LC’s neurological manifestations are not caused by antibody-mediated neuronal injury, with implications for current and future Long COVID treatment trials, which target an autoimmune hypothesis.

## Strengths and Limitations of the study

This study leveraged deeply phenotyped cohorts with a collectively broad set of specimen types, enabling cross-compartment comparisons of humoral immune signals in n-LC. Importantly, neurologic status was assessed using complementary approaches that capture both subjective symptom burden and objective cognitive impairment, reducing reliance on any single readout. Finally, proteome-wide antibody profiling by PhIP-Seq provided an unbiased, high-throughput framework to test whether a shared antibody signature exists across individuals with n-LC.

A key limitation is cohort size, particularly for the Yale COVID Mind study CSF (n=31; n-LC), and may not capture the full biological heterogeneity of n-LC symptoms. However, even within the larger EPICC cohort (n=73), LR achieved only modest discrimination (AUC=0.62) from controls (n=52), suggesting that the observed lack of a shared antibody signature might reflect true biological diversity rather than limited statistical power. Phenotyping also has some limitations, e.g., BRACE as we used it is designed as a self-administered battery of a limited number of standard cognitive tests and thus may lack the granularity needed to precisely define ‘cognitively impaired’ versus ‘un-impaired’, particularly for subtle or domain-specific deficits. In the Yale COVID Mind study, neurologic symptom classification may additionally be influenced by the non-specificity of some symptoms and variability and noise in patient-reported outcomes.

Finally, while PhIP-Seq offers broad, unbiased coverage, the platform primarily detects linear peptide epitopes and can miss conformational, glycosylated, multimeric, or post-translationally modified antigens typical of many neuronal surface autoantibodies and cytokines/chemokines. An important future direction is to pair PhIP-Seq with complementary antigen-presentation formats and assays, such as emerging yeast-based display libraries (e.g., REAP) that can present longer or more structurally informative antigens, to better capture non-linear epitope biology and strengthen mechanistic interpretation.

## Materials and Methods

### Participants and sample collection

#### Sex as a biological variable

Our study cohorts included both female and male patients, as shown in Table 1. Findings are expected to apply to both sexes.

#### Yale COVID Mind Study

The COVID Mind Study at Yale (HIC#1502015318) is a longitudinal, outpatient-based study. Three groups were prospectively enrolled (Figure 1a): (1) 31 individuals with n-LC were enrolled; (2) 7 individuals who recovered from COVID-19 with no LC symptoms were enrolled; and (3) samples from 22 healthy individuals who were recruited before 2020 were utilized. The WHO definition (53) was applied during screening and enrollment, supported by standardized surveys and a comprehensive medical and psychiatric history reviewed by a clinician. n-LC was defined as the development of new neurologic symptoms 3 months after a confirmed SARS-CoV-2 infection (RT-PCR or rapid antigen test), with these symptoms lasting for at least 2 months with no other explanation. n-LC participants were recruited from the Yale NeuroCOVID Clinic, where they were seen and diagnosed by a trained neurologist. Participants underwent clinical assessment (screening, surveys, and chart review), blood and CSF collection within a median of 365 days after acute COVID-19 infection, and a 3-hour neuropsychological testing battery. Exclusion criteria included age <18, pregnancy, major and/or pre-existing neurologic illness, major and/or pre-existing psychiatric disorder, and current or past drug use.

#### IDCRP EPICC Study

The EPICC study is a longitudinal study of COVID-19 outcomes in Military Health System (MHS) beneficiaries. The EPICC study enrolled participants between March 2020 and May 2022 across nine U.S. military treatment facilities and an online recruitment pathway (45, 46). Briefly, eligibility criteria included diagnosis with SARS-CoV-2 or testing for SARS-CoV-2; in 2021, eligibility criteria were expanded to those who received a licensed COVID-19 vaccine. The EPICC study collected demographic and clinical data across one year of follow-up, including acute COVID-19 symptoms and severity, comorbidities, and COVID-19 vaccination history; data sources included participant-filled surveys, case report forms completed by site staff, and electronic medical record data from the MHS Data Repository (MDR). Data collection was augmented by upper respiratory swab and blood specimen collection for SARS-CoV-2 virological testing and host response assays, respectively. SARS-CoV-2 infection status was ascertained by medical record review, SARS-CoV-2 qRT-PCR testing of upper respiratory swabs or clinical residual swabs, or participant self-report.

The EPICC study had several specific modules to perform further focused assessment on COVID-19 outcomes; one of these modules was focused on objective assessment of cognitive impairment after SARS-CoV-2 infection with the use of the Brain-Baseline Assessment of Cognition and Everyday Functioning app on an iPhone or iPad device (BRACE; Digital Artefacts LLC, Iowa City, IA) (54). From August 2022, active EPICC participants were offered enrollment into the BRACE sub-study, with consenting participants accessing the BRACE app downloaded onto Apple iOS® devices for self-testing. The BRACE app has previously been used to evaluate cognition in the context of other infectious diseases (55). BRACE is a screening measure of objective cognitive function and this sub-study used the following BRACE tests: (i) Trails Making Test A (which measures psychomotor speed), (ii) Trails Making Test B (which measures set shifting and executive function); (iii) Stroop (characterized by word, color, and interference tests) to measure speed of information processing and executive function (behavioral inhibition); and (iv) a visuospatial short-term memory task (VSTM) to assess visuospatial learning and memory.

BRACE outcomes were expressed as the average time to complete all components in each task for the Trails Making tests and the Stroop tests. For the VSTM task, BRACE outcomes were reflected as percent accuracy. Values were log-transformed for normalization (for continuous time variables), and longer times reflected lower performance. An adjustment was made for iOS device size (iPhone versus iPad). BRACE impairment was defined as a T-score < 40 (i.e., one standard deviation below the population mean). In addition to adjusting the Trails values based on the platform (iPhone or iPad) on which it was taken, we also estimated the Trails t-scores separately for iPhone and iPad users. This sensitivity analysis resulted in no changes to the BRACE Trails impairment status variables in the dataset.

This BRACE assessment complemented a previous evaluation of self-reported cognitive function after SARS-CoV-2 infection using the PROMIS® Cognitive Function and PROMIS® Cognitive Function abilities tools (56). Of those EPICC participants who completed the BRACE module, a subset of SARS-CoV-2-positive participants was sub-selected for analysis in this autoimmunity study based on the availability of blood samples collected after SARS-CoV-2 infection and before BRACE measurement (Figure 1b).

### Mouse brain tissue-based immunofluorescence

C57BL/6J mice from Jackson Laboratory were perfused between P40 and P80 with 7.5 mL of ice-cold PBS, followed by 7.5 mL of 4% paraformaldehyde. Brains were post-fixed for 2-4 hours on ice, cryoprotected overnight in 15% sucrose in PBS, following another overnight cryoprotection in 30% sucrose in PBS. Brains were embedded in 30% sucrose in OCT and cryosectioned sagittally at 12 um thickness. Sections were stored at -20°C.

To stain, slices were re-hydrated with three 5-minute washes in 0.1% Triton-X 100 in PBS (PBST). Slices were blocked with 10% lab serum in 0.2% Triton-X 100 in PBS for 60-90 minutes. CSF was diluted 1:10 in blocking buffer, and slides were stained in diluted CSF overnight at 4°C. The next day, slides were again washed three times for 5 minutes with PBST, then stained for one hour with Alexa Fluor-conjugated secondary antibodies (Jackson ImmunoResearch, catalog number 709-545-149) diluted 1:500 in blocking buffer. Sections were stained for 5 minutes with a 1:5000 dilution of DAPI in PBST, then washed three more times in PBST for 5 minutes each. ProLong Gold Antifade Mountant (Thermo Fisher, catalog number P36930) was used to mount coverslips. Slides were imaged on a Zeiss Axioscan microscope at 20x magnification. CSF was screened at 1:10; positives were re-stained at 1:4 for pattern interpretation. Images were analyzed compared to a negative control lacking any primary antibodies. Brain sections were assessed in a blinded manner. Two people read the tissue slides and crosschecked the observations.

### Phage Immunoprecipitation Sequencing (PhIP-Seq)

To profile proteome-wide antibody repertoires, the PhIP-Seq protocol was employed. The phage display library consisted of approximately 731,724 synthetic peptides, each 49 amino acids in length and tiled with a 25-amino-acid overlap across the entire human proteome, including all known isoforms (as of 2016). The detailed protocol is available at https://www.protocols.io/view/derisi-lab-phage-immunoprecipitation-sequencing-ph-4r3l229qxl1y/v1. Briefly, deep-well 2ml round-bottom 96-well plates were blocked overnight at 4°C using 3% bovine serum albumin (BSA) in TBST to reduce non-specific binding. Blocking buffer was removed and replaced with 500 µL of the phage library (10^10^ plaque-forming units per mL) along with either 1 µL of human serum or 40 µL of CSF diluted in 1:1 storage buffer (phosphate-buffered saline supplemented with 0.04% sodium azide, 40% glycerol, and 40 mM HEPES) for immunoprecipitation. Plates were incubated overnight at 4°C on a rocking platform to facilitate antibody-phage interactions. Protein A and G magnetic beads (Thermo Fisher Scientific, 10002D and 10004D) were washed in TNP-40 buffer and added to each well to capture immune complexes. After 1 hour of incubation at 4°C, beads were transferred to filter plates compatible with vacuum manifolds and subjected to five sequential washes with RIPA buffer. Between each wash, beads were incubated for five minutes at 4°C with gentle rocking. 150 µL LB-Carb was added to the immunoprecipitated phage and amplified in 550 µL log-phase *Escherichia coli* strain BLT5403 (OD₆₀₀ = 0.3-0.6). After ∼2 hours of shaking at 1200 rpm to allow complete lysis, NaCl was added to a final concentration of 0.5 M to facilitate full lysis. Lysates were centrifuged at 3500 rpm for 1 hour, and the supernatant was transferred to another BSA-blocked round-bottom plate. The supernatant underwent a second round of immunoprecipitation and amplification to enrich specific antibody-bound phage. Final lysates were centrifuged and stored at 4°C for subsequent library preparation. Phage DNA was barcoded and amplified using Phusion PCR (18 cycles) and subjected to next-generation sequencing (NGS) using an Illumina NovaSeq platform, with an average sequencing depth of approximately one million reads per sample. Samples were run in duplicate.

### PhIP-Seq Analysis and Statistics

Data analysis was performed using both R and python programming languages. Raw sequencing reads were trimmed and aligned to the reference PhIP-Seq database using RAPSearch (v2.2). Peptide counts were normalized to reads per 100,000 (rpK) for each sample by dividing each peptide count by the sum and multiplying by 100,000. The resulting peptide rpK counts were used to calculate fold change (FC). First, FC was calculated with respect to the average RPK of negative control protein A/G magnetic beads to identify all peptides that were enriched in samples. Then, FC were calculated with respect to the average rpK of healthy controls to identify peptides that are significantly enriched in cases. Comparisons done between n-LC and healthy controls were computed for CSF and serum separately. For all comparisons, PairSeq (57) was used to identify enriched peptides with the following thresholds: FC_THRESH1 >= 10, FC_THRESH2 >= 100, SUM_RPK_THRESH >= 50, KMER_SIZE = 7. All titin and obscurin peptides were omitted from results. Candidate peptides were then filtered to be enriched in >2 cases relative to healthy controls, enriched across replicates in a least one patient and have a sum of RPK across cases > 100. In addition, p-values were calculated for each peptide (cases compared to controls) and adjusted using the Benjamini-Hochberg (BH) method for multiple hypothesis testing. Only peptides with an BH adjusted p-value < 0.05 were kept. A custom scoring function was used to rank peptides by their enrichment in the group of interest relative to controls. Peptides meeting the thresholds and belonging to a gene found in the top scoring peptides were then included in heatmaps. The heatmaps show log_10_ transformed fold change values, calculated from cases with respect to the average rpK of healthy controls.

### Logistic Regression Analysis

A supervised logistic regression (LR) classifier was generated to evaluate whether PhIP-Seq antibody reactivity profiles could distinguish case-control status across cohorts and to classify cognitive impairment status in the EPICC cohort. An input count matrix of per-peptide rpK values was used across all peptides in the PhIP-Seq library. Only peptides that met the minimum rpK threshold of 500, were considered in the modeling process. For each analysis, one hundred rounds of random sampling without replacement were performed on the filtered rpK matrix. This resulted in 100 combinations of training and test datasets. The training data were used to calculate probability scores which informed the prediction results of the test sets. The collective prediction results across all 100 iterations were visualized using a receiver operating characteristic (ROC) curve. The ROC curve utilized incremental probability thresholds from 0 to 1 in order to evaluate the performance of each model. The area under the curve (AUC) was calculated for all 100 iterations as well as the average ROC curve. Of the 100 iterations, the highest-performing model (with the largest AUC) and the lowest-performing model (with the lowest AUC) were both visualized. In addition, a curve representing the average of all 100 iterations was plotted. Separately, for each model, a table of model coefficients was calculated across peptides using the input training datasets. Peptides with the largest coefficient values contributed the most to the probability estimates of each model. Across all iterations of training data, these coefficient values were averaged for each peptide.

### Split-luciferase binding assay (SLBA)

For orthogonal validation of candidate autoantigens, SLBA was performed as previously described. A detailed protocol is available on protocols.io (https://www.protocols.io/view/split-luciferase-binding-assay-slba-protocol-4r3l27b9pg1y/v1). DNA coding for the desired peptide for use in SLBAs was inserted into split luciferase constructs containing a terminal HiBiT tag and synthesized as DNA oligomers (Twist Biosciences). DNA oligomers were amplified by PCR using the indicated primer pairs (5′-AAGCAGAGCTCGTTTAGTGAACCGTCAGA-3′ and 5′-GGCCGGCCGTTTAAACGCTGATCTT-3′).

The unpurified PCR products served as input for the T7 TNT in vitro transcription/translation kit (L1170, Promega). Relative luciferase units of the translated peptides were quantified using the Nano-Glo HiBiT Lytic Detection System (N3040, Promega) in a luminometer. Equal amounts of translated protein were then incubated overnight at 4°C with 20 µl of patient CSF (diluted in 1× storage buffer) and 1 µl of anti-HiBiT positive control antibody (1:10 dilution; CS2006A01, Promega). Immunoprecipitation was performed on 25 µl of Sephadex Protein A/G beads (1:1 ratio; GE17-5280-02 and GE17-0618-05, Sigma-Aldrich) using 96-well polyvinylidene difluoride filtration plates (EK-680860, Corning). After extensive washing, luminescence was measured using the Nano-Glo HiBiT Lytic Detection System (N3040, Promega) in a luminometer. Each sample was run in duplicate. Antibody index was calculated as follows: (sample value - mean blank value)/ (positive control antibody values - mean blank values). Statistical comparisons between two groups of samples were performed using a two-tailed Mann-Whitney test.

### Cell-Based Overexpression Assay (CBA)

HEK293 cells were plated onto poly-d-lysine-coated coverslips. The following day, lipofectamine 3000 (Thermo Fisher Scientific, L3000015) was used to transfect cells with the plasmid of interest, according to Thermo Fisher’s supplied protocol. Cells were allowed to recover overnight before being rinsed in cold PBS and fixed with 4% PFA for 10 minutes. Cells were rinsed four times with PBS, then blocked and permeabilized with 10% lamb serum and 0.5% Triton X-100 in PBS. CSF was diluted 1:10 in the above blocking solution, along with 1:2000 mouse anti-FLAG antibody (Millipore F3165). The cells were then stained overnight in the diluted CSF and anti-FLAG solution. The next day, cells were washed three times with PBS, then stained for one hour with Alexa Fluor-conjugated secondary antibodies (Jackson ImmunoResearch 715-585-151, 709-545-149) diluted 1:500. Sections were stained for 5 minutes with a 1:5000 dilution of DAPI in PBST, then washed three more times in PBS. ProLong Gold Antifade Mountant (Thermo Fisher P36930) was used to mount coverslips.

### Luminex bead-based assay

To determine whether n-LC reflected an ongoing/persistent anti-viral antibody response in the CNS, SARS-CoV-2 specific antibodies were quantified in paired CSF and serum samples using Luminex bead-based immunoassay in participants with n-LC compared to both control groups in the Yale COVID Mind study (58). Briefly, total IgG concentrations in paired CSF and serum samples from n-LC and both sets of controls were quantified using a commercial ELISA kit (Fortis Life Sciences, Cat# E88-104) according to the manufacturer’s protocol. CSF samples were diluted 1:20 in PBST (PBS + 0.05% Tween-20 with 2% non-fat milk). Serum samples were diluted to match the IgG concentration of the paired CSF. Samples were incubated for 1 hour at room temperature with MagPlex microspheres (10,000,000 beads/mL) coated with S, N, or BSA (negative control), each assigned a unique bead ID. Following washing with PBST, beads were incubated for 30 minutes at room temperature with phycoerythrin-conjugated anti-human IgG (1:2000; Invitrogen, Cat# 12-4998-82). After a final wash, beads were analyzed in 96-well plates on a Luminex LX-200 cytometer. Antibody binding to each antigen and to BSA was reported as median fluorescence intensity (MFI). MFIs corresponding to bead IDs for S and N were extracted and normalized to the BSA MFI for each sample. Antibody index values were derived from the normalized MFIs, calculated as (CSF MFI/Serum MFI) / (CSF total IgG/Serum total IgG), and statistically compared across all groups by the Kruskal-Wallis test.

## Study approval

All studies in the Yale COVID Mind study cohort were approved by the Yale IRB (HIC#1502015318), and written consent was obtained from all participants. All EPICC participants also provided informed consent, and the EPICC protocol was approved by the USU Institutional Review Board (Protocol number- IDCRP-085).

## Data Availability

PhIP-seq data are available for download at Dryad (https://doi.org/10.5061/dryad.kprr4xhkt), and code is deposited in GitHub (https://github.com/UCSF-Wilson-Lab/NeuroLongCOVID_Yale_EPICC_PhIPseq). Values for all data points in graphs are reported in the Supporting Data Values file.

## Author Contributions

DC contributed to conceptualization, data curation, formal analysis, investigation, methodology, project administration, resources, software, supervision, validation, visualization, and writing of the original draft, as well as review and editing of the manuscript. RD, VDL, and IL contributed to data curation, formal analysis, methodology, visualization, and writing of the original draft; RD and VDL also contributed software, and IL additionally contributed to validation. LMA contributed to resources, investigation, funding, methodology, and review and editing of the manuscript. JC, AN, and KZ contributed to data curation, project administration, and resources. TN and PC contributed to data curation and investigation. CYW, AS, and BCR contributed to investigation. DRT contributed to writing, review, and editing. THB, LHR contributed to review and editing of the manuscript. SAR contributed to data curation, methodology, and review and editing of the manuscript. BKA, SDP, SF, SS, and SM contributed to conceptualization, data curation, funding acquisition, project administration, resources, supervision, and review and editing of the manuscript. MRW contributed to conceptualization, data curation, formal analysis, funding acquisition, methodology, project administration, software, resources, supervision, visualization, and writing of the original draft, as well as review and editing of the manuscript.

## Disclaimer

Views expressed are those of the authors and do not reflect the official policy of USUHS, the Department of the Army, Department of the Navy, the Department of the Air Force, the Department of War, the Defense Health Agency, or the U.S. Government, or the Henry M. Jackson Foundation for the Advancement of Military Medicine, Inc. (HJF). The investigators have adhered to the policies for protection of human subjects as prescribed in 45 CFR 46.

## Copyright

Some coauthors are U.S. Government employees or service members and the work described was created as part of their official duties. Title 17 U.S.C. §101 defines a U.S. Government body of work as a work created by an employee of the U.S. Government or military service member as part of that person’s official duties. Title 17 U.S.C. §105 reports “Copyright protection under this title is not available for any work of the United States Government.”

## Supporting information

Supplementary Figures 1 to 6 with their figure legends

## Data Availability

All PhIP-seq data are available for download at Dryad and code is deposited in GitHub.

## Acknowledgements

We thank the patients and their families for their participation in this study. We thank the members of the EPICC COVID-19 Cohort Study Group for their many contributions in conducting the study and ensuring effective protocol operations. The following members were all closely involved with the design, implementation, and oversight of the overall EPICC study.

Brooke Army Medical Center, Fort Sam Houston, TX: J. Cowden; M. Darling; S. DeLeon; D. Lindholm; A. Markelz; K. Mende; S. Merritt; T. Merritt; N. Turner; T. Wellington

Carl R. Darnall Army Medical Center, Fort Hood, TX: S. Bazan; P.K Love

Fort Belvoir Community Hospital, Fort Belvoir, VA: N. Dimascio-Johnson; N. Elnahas; E. Ewers; K. Gallagher; C. Glinn; U. Jarral; D. Jennings; D. Larson; K. Reterstoff; A. Rutt; A. Silva; C. West Henry M. Jackson Foundation, Inc., Bethesda, MD: P. Blair; J. Chenoweth; D. Clark

Madigan Army Medical Center, Joint Base Lewis McChord, WA: J. Bowman; S. Chambers; C. Colombo; R. Colombo; C. Conlon; K. Everson; P. Faestel; T. Ferguson; L. Gordon; S. Grogan; S. Lis; M. Martin; C. Mount; D. Musfeldt; D. Odineal; M. Perreault; W. Robb-McGrath; R. Sainato; C. Schofield; C. Skinner; M. Stein; M. Switzer; M. Timlin; S. Wood

Naval Medical Center Portsmouth, Portsmouth, VA: S. Banks; R. Carpenter; L. Kim; K. Kronmann; T. Lalani; T. Lee; A. Smith; R. Smith; R. Tant; T. Warkentien

Naval Medical Center San Diego, San Diego, CA: C. M. Berjohn; S. Cammarata; N. Kirkland; D. Libraty; R. Maves; G. Utz

Tripler Army Medical Center, Honolulu, HI: C. Bradley; S. Chi; R. Flanagan; A. Fuentes; M. Jones; N. Leslie; C. Lucas; C. Madar; K. Miyasato; C. Uyehara

Uniformed Services University of the Health Sciences, Bethesda, MD: H. Adams; B. Agan; L. Andronescu; A. Austin; B. Barton, D. Becher, C. Broder; T. Burgess; C. Byrne; K Chung; J. Davies; C. English; N. Epsi; C. Fox; M. Fritschlanski; A. Hadley; P. Hickey; E. Laing; C. Lanteri; J. Livezey; A. Malloy; A. Michel, R. Mohammed; C. Morales; P. Nwachukwu; C. Olsen; E. Parmelee; S. Pollett; S. Richard; J. Rothenberg, J. Rozman; J. Rusiecki; D. Saunders; E. Samuels; M. Sanchez; A. Scher; M. Simons; A. Snow; K. Telu; D. Tribble; M. Tso; L. Ulomi; M. Wayman, N. Hockenbury, J. Rothenberg

United States Air Force School of Aerospace Medicine, Dayton, OH: T. Chao; R. Chapleau; M. Christian; A. Fries; C. Harrington; V. Hogan; S. Huntsberger; K. Lanter; E. Macias; J. Meyer; S. Purves; K. Reynolds; J. Rodriguez; C. Starr

United States Coast Guard, Washington, DC: J. Iskander; I. Kamara

Womack Army Medical Center, Fort Bragg, NC: B. Barton; D. Hostler; J. Hostler; K. Lago; C. Maldonado; J. Mehrer

William Beaumont Army Medical Center, El Paso, TX: T. Hunter; J. Mejia; R. Mody; J. Montes; R. Resendez; P. Sandoval

Walter Reed National Military Medical Center, Bethesda, MD: I. Barahona; A. Baya; A. Ganesan; N. Huprikar; B. Johnson

Walter Reed Army Institute of Research, Silver Spring, MD: S. Peel

