## Supplementary Figures 1 to 6 with their figure legends for "Autoantibody landscapes in neurological Long COVID and post-COVID cognitive impairment show heterogeneity without a shared disease signature"

**Supplementary Data:**

**Supplementary Figure 1**

**a) Case_nLC_Y10**

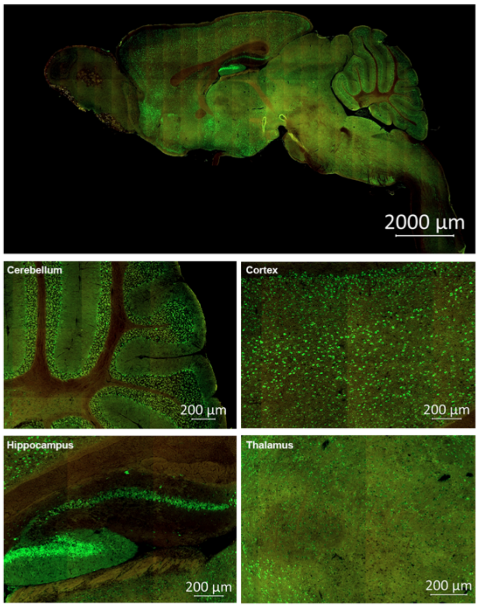


**b) Case_nLC_Y21**

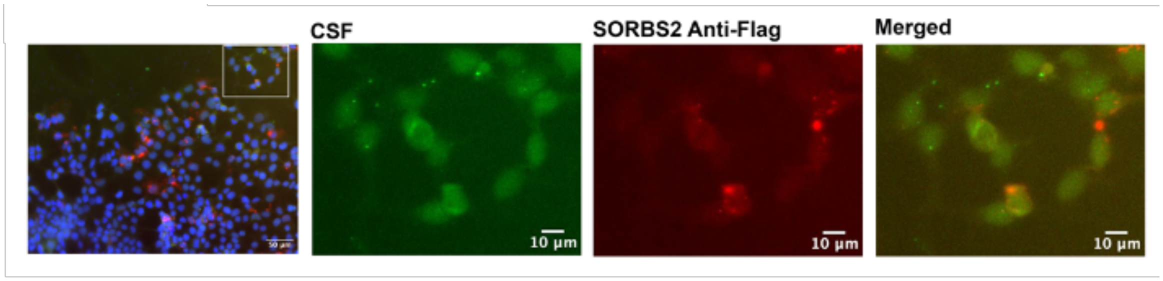


**Supplementary Figure 1. a) Case example showing pronounced neuropil reactivity.** Mouse sagittal brain sections were incubated with patient CSF (1:4) and detected with anti-human IgG (green). The composite whole-brain image (top; scale bar, 2,000 µm) is accompanied by higher-magnification fields (bottom; cerebellum, cortex, hippocampus, thalamus; scale bars, 200 µm). This case exhibited intense cytoplasmic staining in the nucleus accumbens and deep cerebellar nuclei, together with robust neuropil labeling in the dentate gyrus extending along mossy fiber tracts.

**b) Sorbin and SH3 domain containing 2 protein (SORBS2) enriched in nLC patient above both never-COVID and post-COVID controls by PhIP-Seq, did not validate by HEK293 overexpression cell-based assay**. Cells transfected with FLAG-tagged SORBS2 were immunostained with patient CSF (1:10) and a rabbit anti-FLAG antibody. Cells were then counterstained with anti–human IgG 488 (green) and anti–rabbit IgG 594 (red). No colocalization was observed between the patient CSF immunostaining and SORBS2 overexpression in the HEK293 cells.

**Supplementary Figure 2**

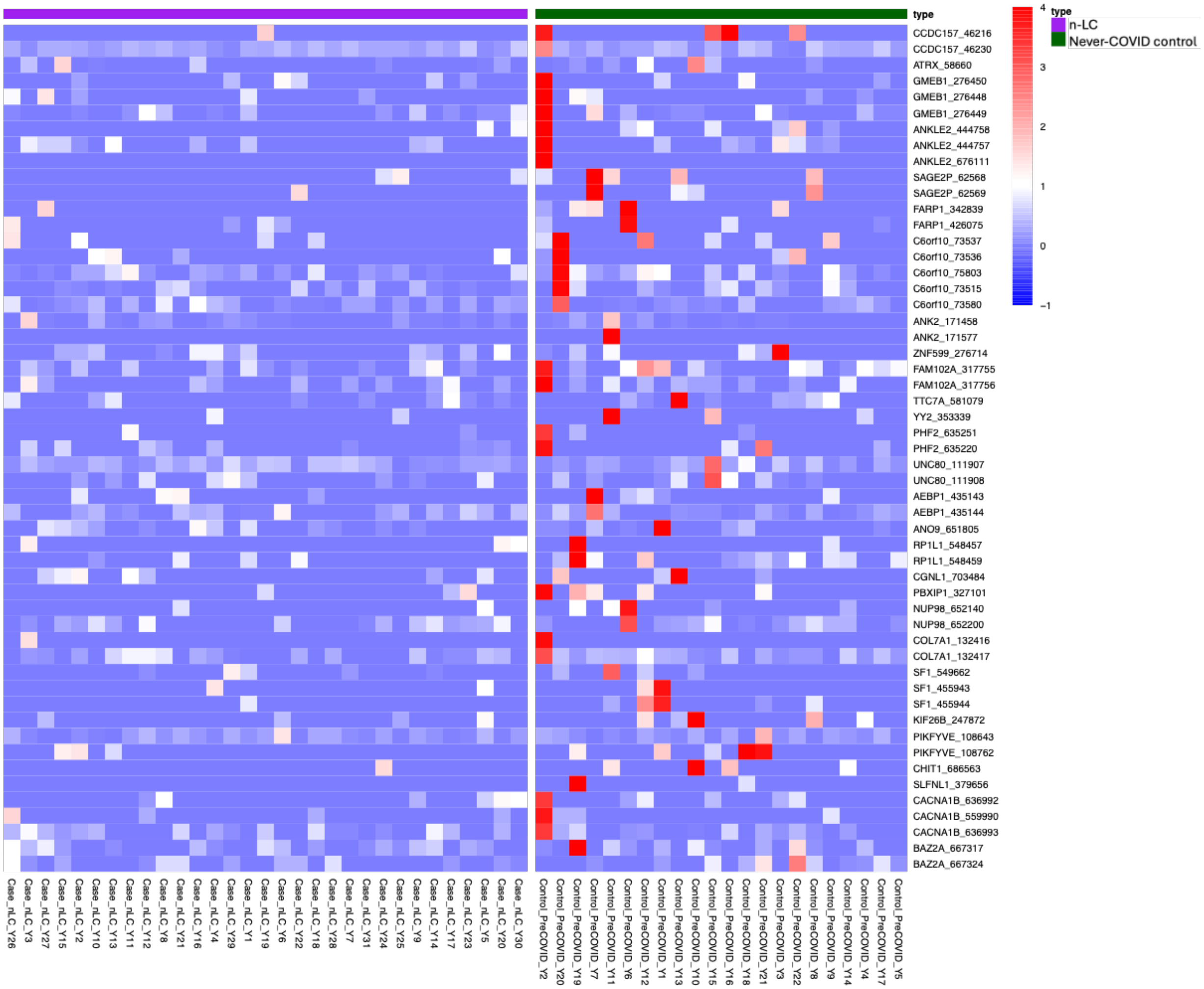


**Supplementary Figure 2. a) Reciprocal analysis of human PhIP-Seq reactivity in Never-COVID CSF controls compared to neurological Long-COVID (n-LC) CSF cases**. Heatmap showing peptide-level enrichment in cerebrospinal fluid (CSF) from Never-COVID control samples relative to patients with neurological Long-COVID (n-LC). Each row corresponds to a unique peptide, and each column represents an individual CSF sample. Color intensity denotes the $log_{10}$-transformed fold change relative to n-LC**.** Peptides were enriched in only 1 or 2 control samples at most, and the peptides enriched in post-COVID asymptomatic controls were distinct from those enriched in n-LC.

**Supplementary Figure 3**

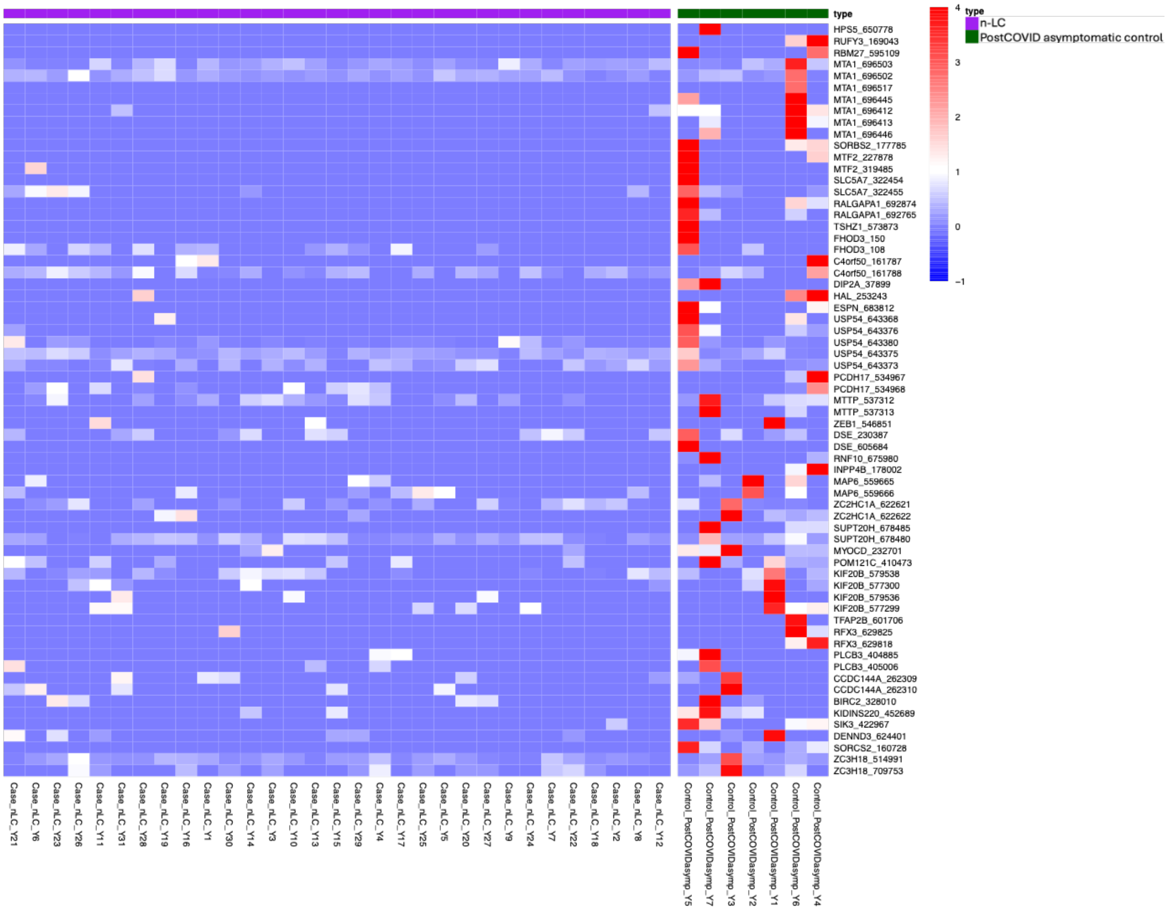


**Supplementary Figure 3. a) Reciprocal analysis of human PhIP-Seq reactivity in post-COVID asymptomatic CSF controls compared to neurological long-COVID (n-LC) CSF cases**. Heatmap showing peptide-level enrichment in cerebrospinal fluid (CSF) from post-COVID asymptomatic control samples relative to patients with neurological Long-COVID (n-LC). Each row corresponds to a unique peptide, and each column represents an individual CSF sample. Color intensity denotes the $log_{10}$-transformed fold change relative to n-LC**.** Peptides were enriched in only 1 or 2 control samples at most, and the peptides enriched in post-COVID asymptomatic controls were distinct from those enriched in n-LC.

**Supplementary Figure 4**

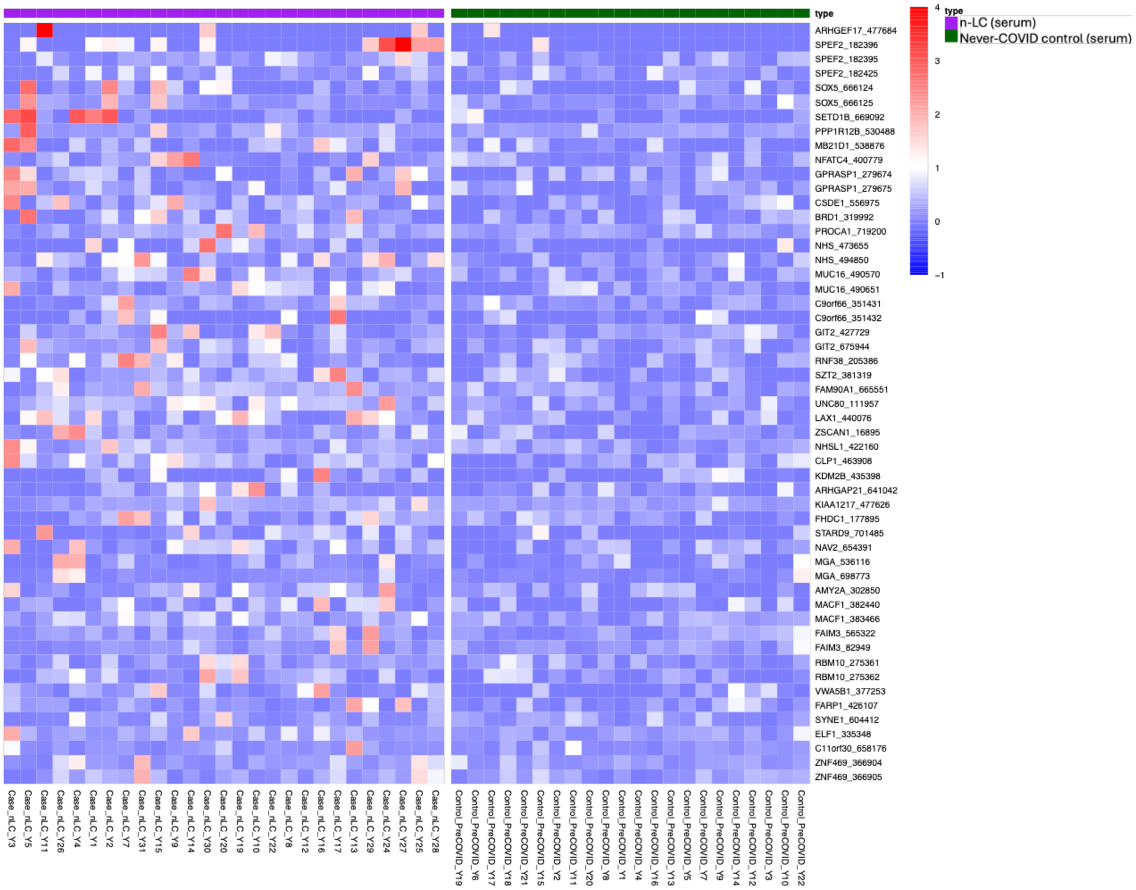


**Supplementary Figure 4. a) Heatmap of human PhIP-Seq reactivity in all n-LC Serum samples compared to Never-COVID controls**. This heatmap displays peptide-level enrichment data from human phage immunoprecipitation sequencing (PhIP-Seq) performed on Serum samples from patients with neurological Long-COVID (n-LC) and pre-pandemic healthy controls (Never-COVID). Each row corresponds to a unique peptide, and each column represents an individual Serum sample. Color intensity reflects the degree of peptide enrichment, quantified as the $\log_{10}$-transformed fold changes in cases relative to controls. Peptides were enriched in only 2-4 patients out of the total n-LC Serum samples above never-COVID controls.

**Supplementary Figure 5**

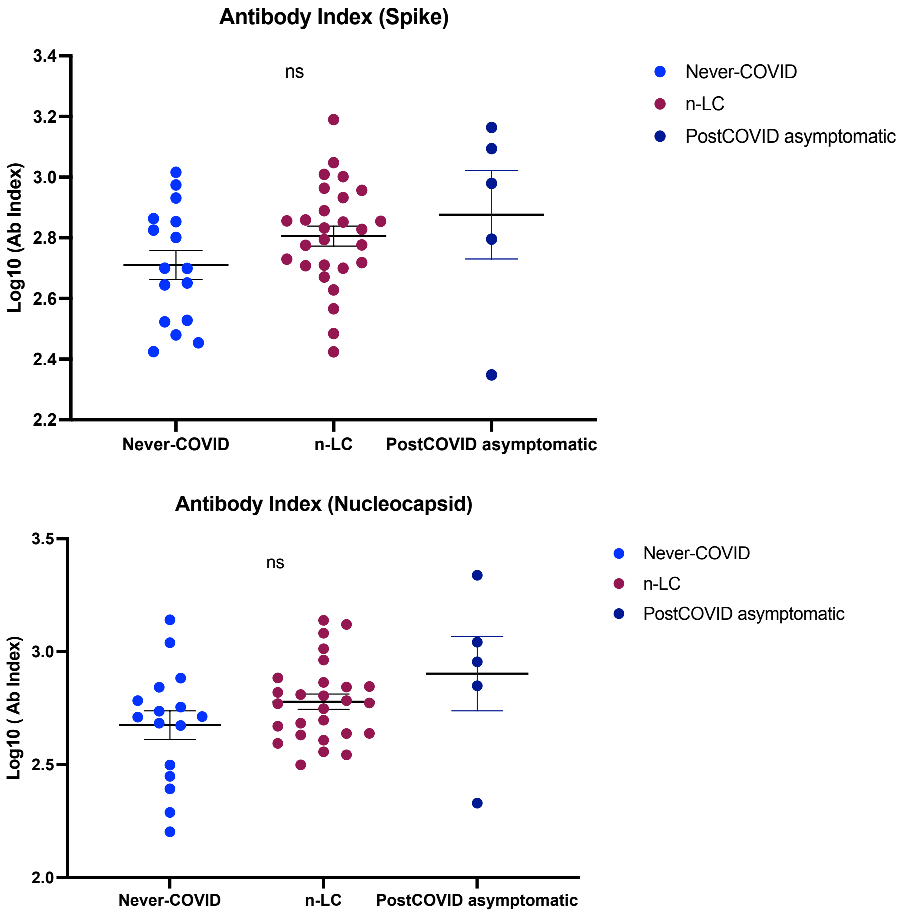


**Supplementary Figure 5. No differences in the SARS-CoV-2 antibody levels in CSF and serum between neurological Long-COVID (n-LC) patients and controls.** A Luminex bead-based immunoassay was used to quantify antibodies against SARS-CoV-2 Spike and Nucleocapsid proteins in CSF and serum. Antibody Index values were calculated as (CSF MFI/Serum MFI) / (CSF total IgG/Serum total IgG). Comparisons among n-LC patients, post-COVID asymptomatic controls, and Never-COVID controls showed no significant differences in the Antibody Index (Kruskal-Wallis test; ns = not significant).

### Supplementary Figure 6

a)

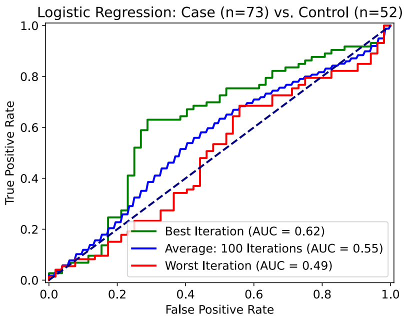


b)

| Top 10 Genes | Logistic Regression Coefficient |
|---|---|
| **NEO1** | 0.10819621070395200 |
| **NEB** | 0.09674775497141580 |
| **ANK2** | 0.08886223796935800 |
| **USP54** | 0.08385534148645630 |
| **TNKS2** | 0.0813342528329224 |
| **NHSL1** | 0.07631398926690000 |
| **LIN9** | 0.06901899177839200 |
| **KIAA1841** | 0.06588981584047080 |
| **SH2D4A** | 0.06546155114152970 |
| **ARID1B** | 0.064846529689791 |

c)

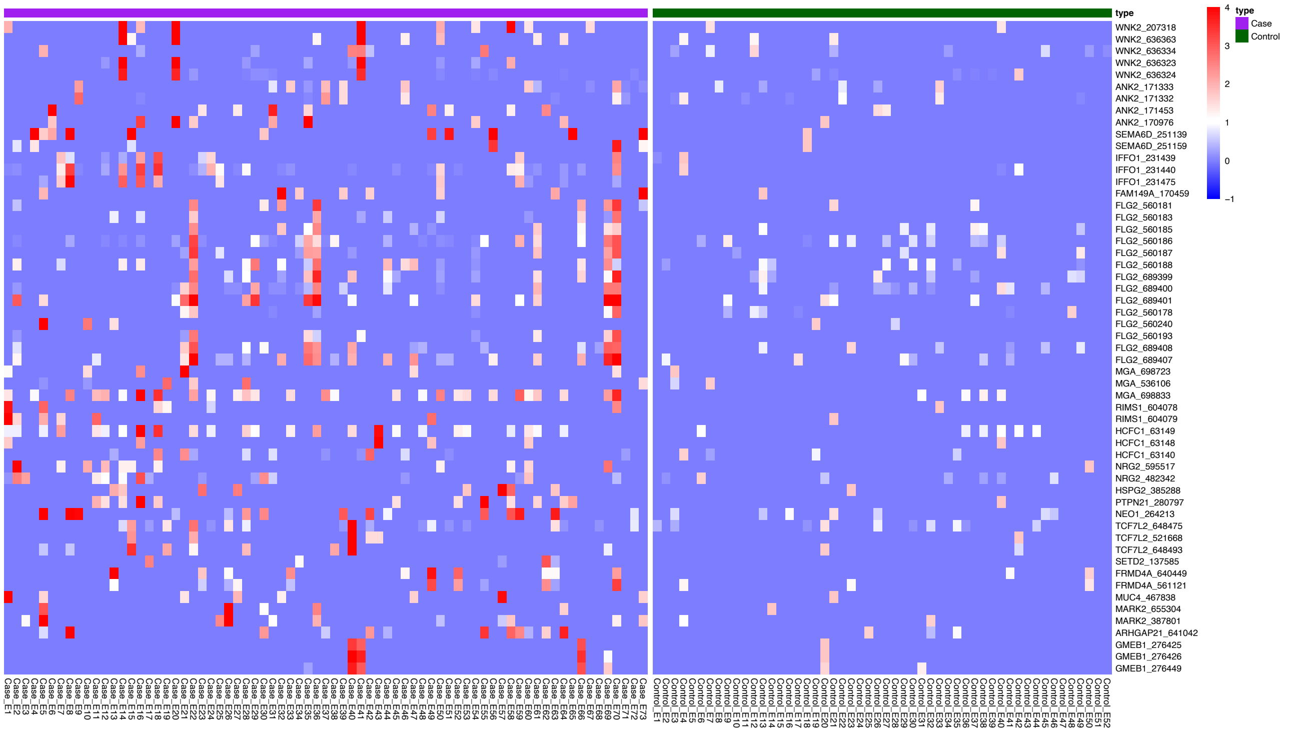


**Supplementary Figure 6. a) Logistic regression classifier only moderately discriminates between all post-COVID serum cases (with or without cognitive impairment) (n = 73) and controls (n = 52) from the EPICC cohort.** ROC curves from a logistic regression classifier trained on peptide-level human PhIP-Seq enrichment data to distinguish post-COVID serum cases (n = 73) from controls (n = 52) across 100 iterations. Discrimination was modest overall, with the best model reaching AUC = 0.62, the mean AUC = 0.55, and the worst iteration AUC = 0.49, indicating only limited predictive performance. **b) Table showing the top genes ranked by their logistic regression coefficients.** These genes contribute the most to case-control classification. **c) Heatmap of human PhIP-Seq reactivity in post-COVID serum cases compared to controls.** This heatmap displays peptide-level enrichment data from human phage immunoprecipitation sequencing (PhIP-Seq) performed on serum samples from post-COVID patients with or without cognitive impairment above controls. Each row corresponds to a unique peptide, and each column represents an individual serum sample. Color intensity reflects the degree of peptide enrichment, quantified as the $log_{10}$-transformed fold changes in cases relative to controls.
